# Single cell profiling of COVID-19 patients: an international data resource from multiple tissues

**DOI:** 10.1101/2020.11.20.20227355

**Authors:** Chan Zuckerberg Initiative Single-Cell COVID-19 Consortia, Esteban Ballestar, Donna L. Farber, Sarah Glover, Bruce Horwitz, Kerstin Meyer, Marko Nikolić, Jose Ordovas-Montanes, Peter Sims, Alex Shalek, Niels Vandamme, Linos Vandekerckhove, Roser Vento-Tormo, Alexandra Chloe Villani

## Abstract

In late 2019 and through 2020, the COVID-19 pandemic swept the world, presenting both scientific and medical challenges associated with understanding and treating a previously unknown disease. To help address the need for great understanding of COVID-19, the scientific community mobilized and banded together rapidly to characterize SARS-CoV-2 infection, pathogenesis and its distinct disease trajectories. The urgency of COVID-19 provided a pressing use-case for leveraging relatively new tools, technologies, and nascent collaborative networks. Single-cell biology is one such example that has emerged over the last decade as a powerful approach that provides unprecedented resolution to the cellular and molecular underpinnings of biological processes. Early foundational work within the single-cell community, including the Human Cell Atlas, utilized published and unpublished data to characterize the putative target cells of SARS-CoV-2 sampled from diverse organs based on expression of the viral receptor ACE2 and associated entry factors TMPRSS2 and CTSL (Muus et al., 2020; Sungnak et al., 2020; Ziegler et al., 2020). This initial characterization of reference data provided an important foundation for framing infection and pathology in the airway as well as other organs. However, initial community analysis was limited to samples derived from uninfected donors and other previously-sampled disease indications. This report provides an overview of a single-cell data resource derived from samples from COVID-19 patients along with initial observations and guidance on data reuse and exploration.

## 1. Introduction & Overview

In late 2019 and through 2020, the COVID-19 pandemic swept the world, presenting both scientific and medical challenges associated with understanding and treating a previously unknown disease. To help address the need for great understanding of COVID-19, the scientific community mobilized and banded together rapidly to characterize SARS-CoV-2 infection, pathogenesis and its distinct disease trajectories. The urgency of COVID-19 provided a pressing use-case for leveraging relatively new tools, technologies, and nascent collaborative networks. Single-cell biology is one such example that has emerged over the last decade as a powerful approach that provides unprecedented resolution to the cellular and molecular underpinnings of biological processes. Early foundational work within the single-cell community, including the Human Cell Atlas, utilized published and unpublished data to characterize the putative target cells of SARS-CoV-2 sampled from diverse organs based on expression of the viral receptor *ACE2* and associated entry factors *TMPRSS2* and *CTSL (Muus et al., 2020; Sungnak et al., 2020; Ziegler et al., 2020)*. This initial characterization of reference data provided an important foundation for framing infection and pathology in the airway as well as other organs. However, initial community analysis was limited to samples derived from uninfected donors and other previously-sampled disease indications.

Initial analysis of viral entry factors in existing single-cell datasets spurred a widespread interest in extending analysis to include data generated from COVID-19 patient samples. There was a need for rapid data generation and sharing to clarify COVID-19 pathophysiology and direct future work. In response, the Chan Zuckerberg Initiative solicited brief proposals from members of their existing communities who have pioneered various aspects of single-cell biology. Proposals were prioritized based on: 1. a demonstrated ability to actively recruit necessary participants, 2. sampling of tissues involved in primary infection or those that appear to be subsequently impacted in COVID-19 pathology, 3. an ability to rapidly generate data, and 4. a willingness to share the data extraordinarily quickly. The teams span clinical and research scientists across a diversity of fields and institutions and are organized below by their main affiliations (Table 1), with full author lists (See Author Table provided as an appendix) including all participating members.

**Table 1:**
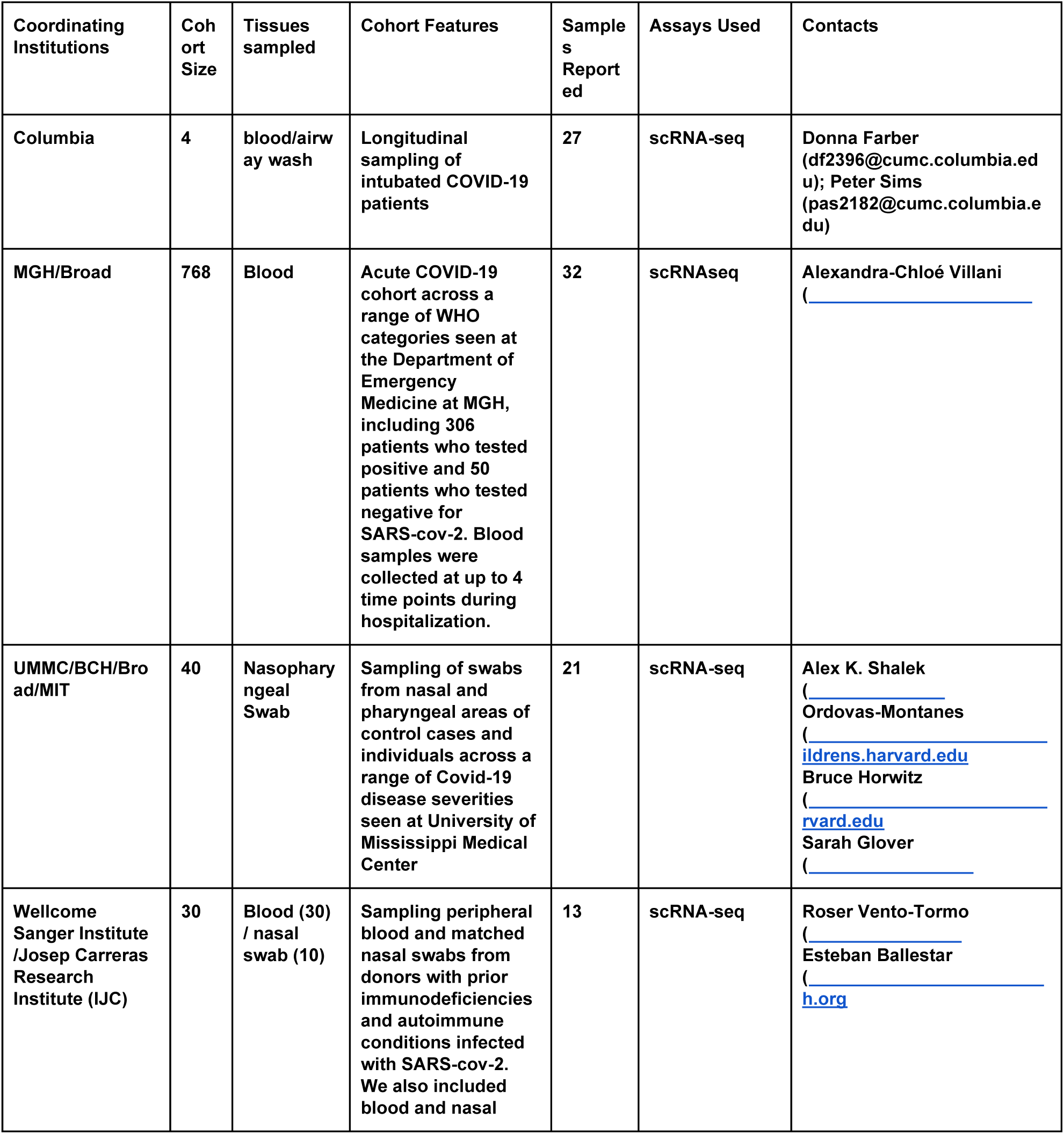
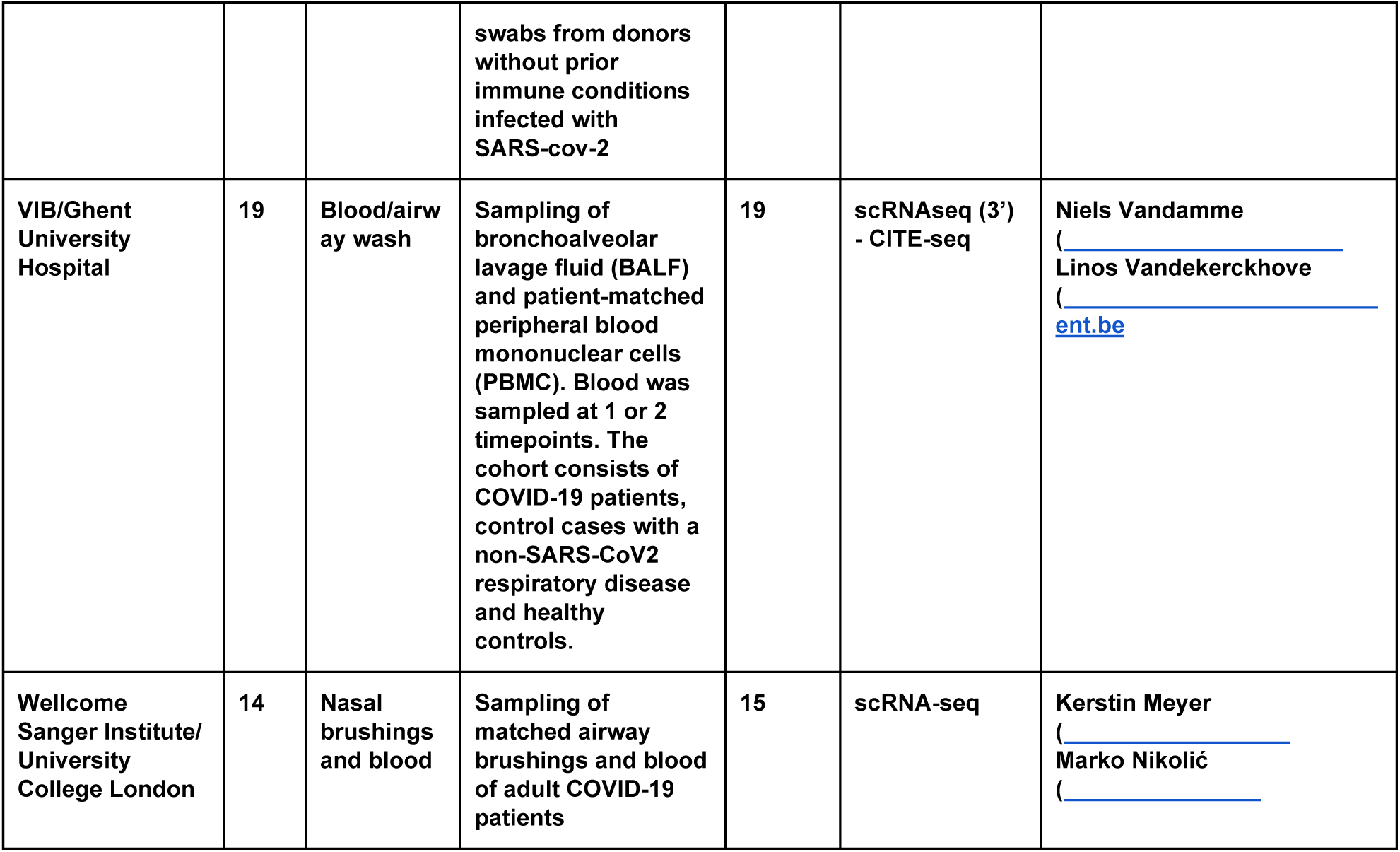
Overview of cohorts and data sets.

All projects aimed to generate and openly share initial data within ∼2 months as a criteria of these awards. Work was initiated in May, 2020, and the teams quickly recruited, processed and analyzed samples allowing for the initial release described here. The pace of these projects was intended to enable data analysis and exploration by other groups, and open possibilities for accelerated use by the scientific community for a variety of critical questions.

A preliminary summary of the cohorts, approaches and data from these initial projects are described below. All teams involved share a goal of providing early data access, recognizing that their own characterizations of these data are works-in-progress and should be treated as such. Further sample collection, sequencing, and analysis is likely to refine what is presented, and we are committed to providing updates as this progresses. Nevertheless, this first release provides a glimpse into: 1. dense temporal sampling of COVID-19 progression; 2. the impact of underlying immune conditions including primary immunodeficiencies and autoimmune diseases; the host response in underrepresented, but disproportionately impacted, participants; 4. age-dependent responses; 4. CITE-Seq analysis; 5. the impact of severe disease across organs; and, 7. sampling of a large cohort. In all cases, the main study limitation is that no dataset is definitive at this stage -- that is, both locked or powered to reveal correlates. However, all data currently collected and preliminarily annotated is being shared now in-line with the goals of the group and to solicit input, provide a community resource, and accelerate our collective understanding of COVID-19. Over time, we hope that these data sets, together with others’, allows the scientific community to answer initial questions such as: 1) What are the *in vivo* cellular targets of SARS-CoV-2 in respiratory tissue? 2) How do cells of the airways and other affected tissues respond to the virus? and, 3) How does the immune system respond over the course of the infection and does this change with the age of the patient and disease severity?

All data can be openly and freely explored and re-used by accessing the data portal at covid19cellatlas.org and clicking on ‘Patient Donors’, though certain stipulations, designed to protect those who worked tirelessly to generate these data, apply (please see Community Standards and Publication Policy). This preprint along with the data and analysis will be updated with additional samples and insight as they emerge in the months to come.

Re-use of this resource is strongly encouraged with the caveat that both the dataset and its interpretation are in a nascent state. This release represents a fraction of the ultimate complete resource, and changes in the diversity of cell types – particularly detection of rare cell subsets – are expected following creation and reanalysis of the larger dataset. Further, the results from preliminary analyses of SARS-CoV-2 viral transcript alignments may change as more robust and conservative statistical procedures are applied to correct for possible confounders. Some groups have provided preliminary cell type annotations, and these may also change as we learn more about this disease and with additional input from additional domain experts. Many cell populations are in highly activated states, which can obscure their identities in comparison to more homeostatic conditions. In many cases, the specimens from these studies present unique challenges for scRNA-seq. Procurement procedures were often suboptimal or difficult to control under the circumstances of a clinical environment during a pandemic, and the disease pathology often presented unexpected complexities. For example, in the Columbia study on severe COVID-19, both the blood and airway samples contained extremely high levels of neutrophils, which had to be removed prior to analysis. Samples could also contain higher levels of ambient RNA and multiplets than in other settings. Finally, we urge caution when performing new analyses using these data, paying particular attention to potential cofounders associated with sample collection, which presented unique challenges during a pandemic, and power limitations for uncovering the impact of any one correlate of disease severity (e.g., age, time since infection, gender) given multifactorial nature of COVID-19.

## 2. Community Standards and Publication Policy

Our consortium has publicly released several single-cell genomic datasets and accompanying clinical metadata with the hope that other scientists will immediately use this resource to advance our understanding of COVID-19. Currently, none of the consortium members have independently, or collectively, published any findings describing the presented data in a peer-reviewed journal. At this time, we ask that anyone who intends to publish observations derived directly or indirectly from this resource please contact the consortium members who generated the relevant data prior to submission (Table 1). While we have made every effort to clarify sample information, experimental and analytical methods, and caveats associated with these data (see “Words of Caution” above and below), direct communication with consortium members will prevent misunderstandings or misinterpretations that could lead to scientific errors. In some cases, consortium members may have additional data that were generated outside of the scope of their CZI-funded project, which could also be useful and shareable. Finally, we would like to ensure that all individuals who generated these data are properly credited for their efforts.

## 3. Outline of Cohorts and Study Design

## 4. Summary of Individual Studies & Observations

### 4.1. Columbia University Medical Center

#### Study Overview and Design

We profiled matching blood and airway wash samples for four patients who had been intubated at Columbia University Irving Medical Center for acute respiratory distress syndrome, a serious complication of SARS-CoV-2 infection. We enrolled patients in the medical ICU and satellite ICUs that were established to accommodate the large patient volume under IRB protocol AAAS9659. Enrollment criteria include patients with diagnosed COVID-19 (positive PCR test for SARS-CoV-2) who were intubated during the previous 24-48 hours and exclude patients treated with immunosuppressants or with known immunodeficiencies or cancer. For three of the four patients (COV022, COV026, and COV028), we acquired samples at multiple time points following intubation (**Table 2**). One patient (COV027) died following the first day of intubation. We isolated mononuclear cells from all of these samples and profiled them with scRNA-seq.

**Table 2:**
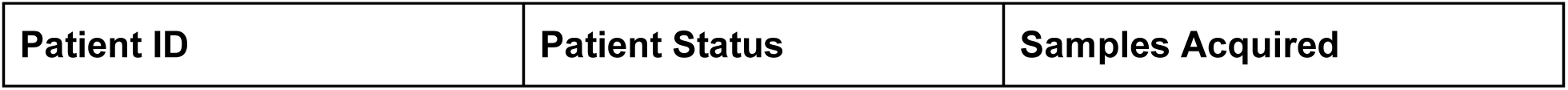
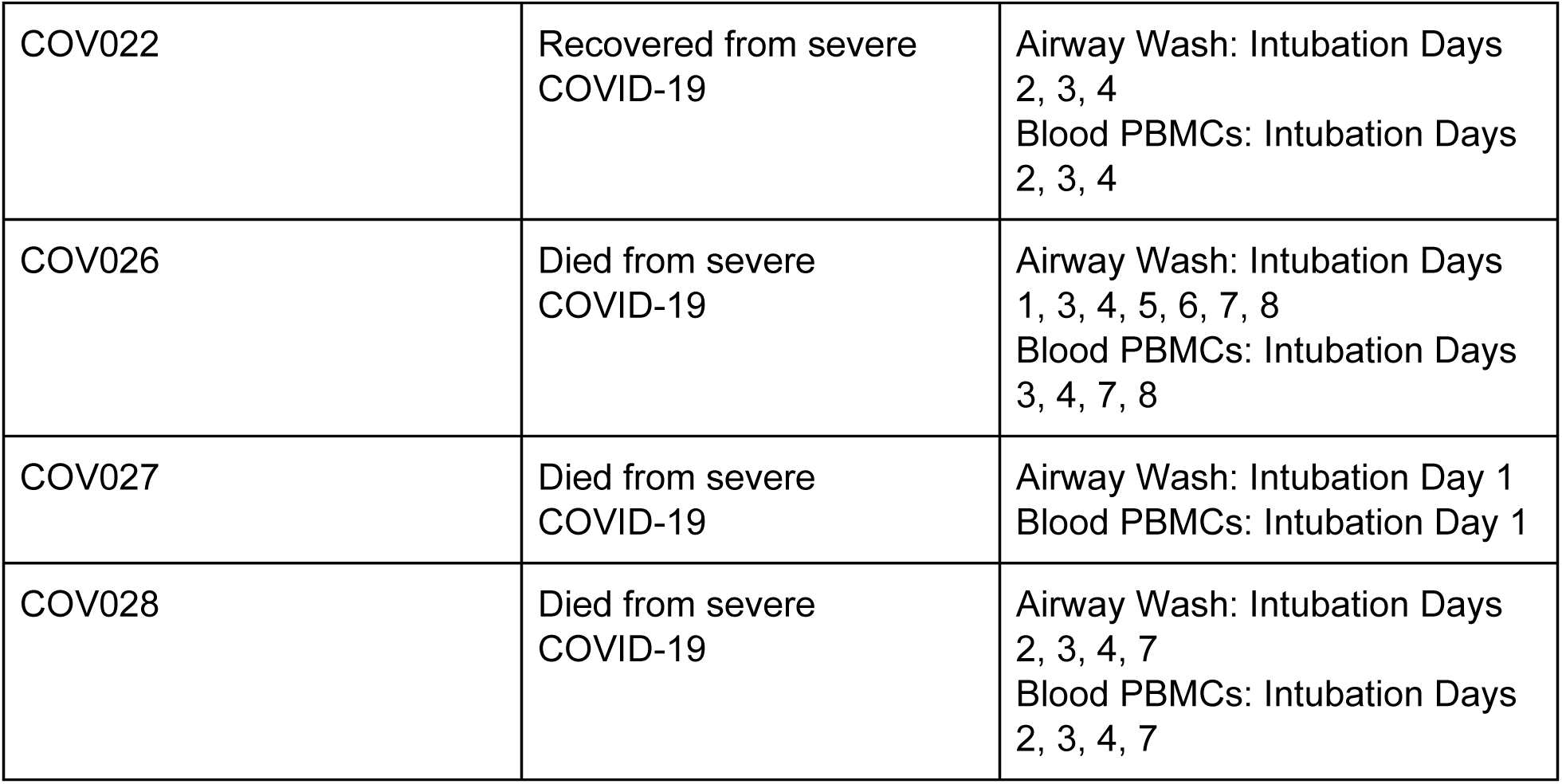
scRNA-seq data sets from Columbia study.

| Patient ID | Patient Status | Samples Acquired |
| --- | --- | --- |
| COV022 | Recovered from severe COVID-19 | Airway Wash: Intubation Days 2, 3, 4<br>Blood PBMCs: Intubation Days 2, 3, 4 |
| COV026 | Died from severe COVID-19 | Airway Wash: Intubation Days 1, 3, 4, 5, 6, 7, 8<br>Blood PBMCs: Intubation Days 3, 4, 7, 8 |
| COV027 | Died from severe COVID-19 | Airway Wash: Intubation Day 1<br>Blood PBMCs: Intubation Day 1 |
| COV028 | Died from severe COVID-19 | Airway Wash: Intubation Days 2, 3, 4, 7<br>Blood PBMCs: Intubation Days 2, 3, 4, 7 |

**Table 3:** Metadata summary for MGH COVID-19 acute blood cohort dataset.

| Meta-data identifier | Category description | Legend annotation |
| --- | --- | --- |
| Time point | Time point at which blood was collected | D0=day visiting the ER; D3=day 3 of hospitalization; D7=day7 of hospitalization; DE=event-driven |
| COVID | COVID status: tested positive prior to enrollment /during hospitalization | 0=negative; 1=positive |
| Age cat | Age category | 1=20-34; 2=36-49; 3=50-64; 4=65-79; 5=80+ |
| BMI cat | Body mass index categories | 0=<18.5 (underweight); 1=18.5-24.9 (normal); 2=25.0-29.9 (overweight); 3=30.0-39.9 (obese); 4 = >=40 (severely obese); 5=Unknown |
| HEART | Pre-existing heart disease: coronary artery disease, congestive heart failure, valvular disease | 0=No; 1=Yes |
| LUNG | Pre-existing lung disease: asthma, COPD, requiring home O2, any chronic lung condition | 0=No; 1=Yes |
| KIDNEY | Pre-existing kidney disease: chronic kidney disease, baseline creatinine >1.5, ESRD) | 0=No; 1=Yes |
| DIABETES | Pre-existing diabetes: pre-diabetes, insulin and non-insulin dependent diabetes | 0=No; 1=Yes |
| HTN | Pre-existing hypertension | 0=No; 1=Yes |
| IMMUNO | Pre-existing immunocompromised condition: active cancer, chemotherapy, transplant, immunosuppressant agents | 0=No; 1=Yes |
| Symp_Resp | Respiratory symptoms: sore throat, congestion, productive or dry cough, shortness of breath or hypoxia, or chest pain | 0=No; 1=Yes |
| Fever_symptom | Fever symptom | 0=No; 1=Yes |
| GI_Symp | Any GI related symptoms at presentation: (abdominal pain, nausea, vomiting, diarrhea) | 0=No; 1=Yes |
| WHO 0 | WHO score for day 0 study window - enrollment plus 24 hours - highest Acuity within Day 0 window | 1 = Death; 2 = Intubated / ventilated, survived; 4 = Hospitalized, supplementary O2 required, survived; 5 = Hospitalized, no supplementary O2 required, survived; 6 = Discharged / Not hospitalized, survived |
| WHO 3 | WHO score for day 3 study window | same as WHO 0 legend |
| WHO 7 | WHO score for day 7 study window | same as WHO 0 legend |
| WHO 28 | WHO score on study day 28 | same as WHO 0 legend |
| WHO max | WHO max is the highest Acuity level between Day 0 -28 | same as WHO 0 legend |
| Trop_72h | Cardiac event: hs-cTn =>100 within first 72 hours of presentation | 0=No; 1=Yes |

#### Sample Collection

We collected paired endotracheal tube (ETT) washes obtained during daily flushing as part of clinical treatment and blood samples daily from the time of enrollment until extubation (recovery) or death. We profiled a subset of these samples using scRNA-seq.

#### Initial Observations and Challenges

To date, we have focused on compositional analysis of matching blood and airway wash specimens from intubated patients. The mononuclear cells isolated from peripheral blood contain myeloid cells, T cells, B cells, plasma cells, neutrophils and dendritic cells. The MNCs for airway washes include myeloid cells, epithelial cells, T cells, B cells, mast cells, ionocytes, and plasmablasts/plasma cells. We observed low levels red blood cell and platelet contamination in a subset of samples from both sources. Importantly, prior to depletion, all of the samples contained very high levels of neutrophils, which we had to remove in order to obtain high-quality scRNA-seq data that were not dominated by a single cell type. In addition, neutrophils have a tendency to lyse prematurely, releasing nucleases and proteases that can compromise mRNA capture and library construction.

Myeloid cells in both the blood and airway were notably diverse both in terms of cellular identity and chemokine expression. Although not included in our original data release, efforts are underway to provide a more comprehensive annotation of myeloid subsets in these specimens. Additionally, we observed a broad diversity of epithelial cells including club, goblet, ionocytes, and likely tuft cells. However, we did not detect reads aligning to the SARS-CoV-2 transcriptome in these or any cells in this data set. Finally, we also obtained good coverage of T cell subsets in both the blood and airway, and based on markers like *ITGA1* and *CXCR6*, the airway washes contain high levels of tissue resident memory (TRM) T cells.

### 4.2 Sanger & IJC

#### Study Overview and Design

Human blood samples were obtained from a cohort of patients under SARS-CoV-2 infection with pre-existing immunological condition, including patients with autoimmune diseases such as rheumatoid arthritis (RA), psoriasis (Ps), Sjogren syndrome (Sj), eosinophilic granulomatosis with polyangiitis (EGPA) and multiple sclerosis (MS). Immunodeficient patients consisted in individuals diagnosed with common variable immunodeficiency (CVID), missing IgA and IgM isotypes and displaying missing or reduced IgG, as well as one individual with lymphopenia and another with bone marrow failure of unknown origin. In some cases, PBMCs samples before, during and after SARS-CoV-2 infection were collected from the same individual (CVID patient 1 and CVID patient 2). For some individuals, nasal swab samples were also collected in parallel with the PBMC sample. For nasal sample collection, the swab was gently inserted along the nasal septum, just above the floor of the nasal passage, to the nasopharynx, until resistance was felt and then the swab was rotated 5 times. The patients were previously diagnosed according to European Society for Immunodeficiencies (ESID) criteria and established criteria for autoimmune diseases (*Perricone and Valesini, 2014*). They were collected at Hospital La Paz, Hospital La Princesa, Hospital Vall d’Hebron, Hospital Can Ruti and Hospital Bellvitge (Spain), and Medical Center-University of Freiburg (Germany). All donors received oral and written information about the possibility that their blood and nasal biopsies would be used for research purposes, and any questions that arose were then answered. Before giving their first sample the donors signed a consent form approved by the Ethics Committee at their corresponding hospitals, which adhered to the principles set out in the WMA Declaration of Helsinki.

#### Initial Observations

Our preliminary analysis of peripheral blood has revealed distinct populations of T cells: CD4+ naive T cells (T4naive: CD4, CD45RA and CCR7); memory CD4+ T cells (T4mem: CD4, CD45RO and CCL5); naive CD8+ T cells (T8naive: CD8, CD45RA and CCR7); memory CD8 T cells (T8mem: CD8, CD45RO and CCL5); regulatory T cells (T4reg: annotated by FOXP3 and IL2RA); and gamma-delta T cells (Tgd: TRDV2). B cells subsets captured included naïve B cells (Bnaive: TCL1A); memory B cells (Bmem: LSP1); and plasma cells (PC: JCHAIN and MZB1). Additionally, we identified two subsets of NK cells (NK1: FCGR3A, CCL5, GZMH; NK2: FCGR3Aneg, CCL5neg, XCL1). The myeloid fraction was comprised by three subsets of monocytes (Classical: CD14+ FCGR3A-; intermediate: CD14+ FCGR3A+; non-classical: CD14low FCGR3A); neutrophils (Neu: FCGR3B); conventional dendritic cells (cDC1: IRF4, CLEC9A; cDC2: CD1C); and, plasmacytoid dendritic cells (pDC: CLEC4C and TNFRSF21). Additionally, we identified erythrocytes (Ery: HBB); platelets (Plt: PPBP, TUBB1 and PF4); and, hematopoietic precursor cells (HSC: CD34).

Several clusters of epithelial cells were defined in the nasal swabs: Squamous cells (SCEL and TMPRSS11E); secretory cells (XBP1 and MUC5AC); and ciliated cells (PIFO, FOXJ1 and MLF1). Immune cells found in the nasal swabs included myeloid, B and T cells.

### 4.3 University of Mississippi Medical Center, Boston Children’s Hospital & Broad Institute

#### Study Overview and Design

Our group is using single-cell RNA-seq (scRNA-seq) to characterize the direct cellular targets of viral infection and bystander cell responses among the nasal mucosa of an adult cohort following COVID-19 diagnosis and inpatient hospitalization at the University of Mississippi Medical Center. Nasopharyngeal swabs are being collected and banked at the patient bedside and then processed for scRNA-seq. To date, we have generated a dataset which includes 21 participants (15 diagnosed with COVID-19, 6 controls; eventual cohort size of 40+ participants). We hypothesize that the intrinsic response of direct viral targets following infection, as well as their interactions with innate and adaptive immune cells and the tissue parenchyma in the nasal mucosa, plays a crucial role in limiting or permitting COVID-19 disease progression and severity. Identifying whether and how upper airway host-virus interactions specify divergent outcomes is essential to understand SARS-CoV-2 pathogenesis and identify treatments and vaccine strategies uniquely suited to individuals suffering from, or at risk for, COVID-19.

Participants aged 18 years and older were recruited and prospectively enrolled by the study team at University of Mississippi Medical Center (UMMC) (Jackson, Mississippi) between April 2020 and July 2020. Informed consent was obtained from the participation or their legally authorized representative in accordance with UMMC IRB#2020-0065. COVID-19 positive participants were recruited from the hospital or acute respiratory clinic. COVID-19 negative participants were recruited from the UMMC GI Endoscopy lab. All individuals undergoing endoscopy were required to have a negative test for SARS-CoV-2. Inclusion criteria for COVID-19 participants included fever, cough, sore throat and/or shortness of breath with presumed diagnosis of COVID-19 upper respiratory tract infection. The patients all weighed 110 lbs or greater. Non-COVID-19 (control) participants all had a negative COVID-19 test, weighed 110 pounds or greater, and were seen in either GI Endoscopy or UMMC Acute Respiratory Clinic. Exclusion criteria for both cohorts included a history of blood transfusion within 4 weeks and subjects who could not be assigned a definitive COVID-19 diagnosis from either nucleic acid testing or Chest CT imaging. For the nasopharyngeal (NP) samples, both genders were represented – males (n=7) and females (n=8). 6 of the participants were non-COVID-19 (control) – 2 identified as male, 4 as female. The median age of patients that provided NP samples was 56 years old. The median age of patients that provided NP samples was 56 years old. The average day at which NP samples were collected ranged from Day 1 to Day 3 of hospitalization. It should be noted that collection day does not uniformly correspond to days post infection or the presentation of symptoms and is a challenge when considering and comparing COVID-19 progression in many studies. The Institutional Review Board approved the study, and all subjects provided written informed consent, or their legally authorized representative provided it on their behalf. Research samples were collected from volunteers in the form of nasal swabs. A healthcare provider collected the nasopharyngeal sample using a single FLOQswab (COPAN, cat# 0123) which was then viably frozen prior to dissociation and scRNA-seq using Seq-Well S^3 (Summary of Methods).

### Initial Observations

The current interim dataset consists of cells derived from 21 study participants: 6 were non-COVID19 SARS-CoV-2 negative controls and 15 were diagnosed with COVID-19 based on SARS-CoV-2 PCR from nasopharyngeal swab sample and chest CT. As of 7/31/20, the dataset consists of 32,389 genes and 12,427 cells, with an additional 27 distinct genomic features from SARS-CoV-2. Cellular recovery across distinct nasal swabs was highly variable, and we recovered slightly fewer cells among nasal swabs from COVID-19 participants, with 499 +/- 145 (mean +/- SEM) cells per participant, compared to 823 +/- 224 cells per control participant (n.s., p = 0.25 by student’s t-test). We speculate that lower cell recovery may derive from higher proportions of dead and dying cells in infected and inflamed samples. We found roughly equivalent quality of cells (following preprocessing steps outlined above) across nasal swabs from COVID19 and control participants.

Following the dimensionality reduction and clustering approaches discussed above, we annotated 21 clusters corresponding to distinct cell types and states across immune and epithelial identities (**Figure 1, Supplementary Table 1:** marker gene lists using ‘bimod’ likelihood-ratio test). As tissue sampling relied on surface-resident cells that were gently scraped off of the nasopharyngeal epithelium, we did not expect to recover stromal cells such as endothelial cells, fibroblasts, or pericytes, which were found in previous scRNA-seq datasets from nasal epithelial surgical samples (*Ordovas-Montanes et al., 2018; Garcia et al., 2019*). We recovered several immune cell populations, including T cells, dendritic cells, and macrophages, but did not find clusters of cells corresponding to plasmablasts, neutrophils, mast cells, and eosinophils, which have been recovered in superficial nasal wash or nasal scraping samples by other groups (*Cao et al., 2020; Ordovas-Montanes et al., 2018*). We suspect further iterations of this dataset with larger sample sizes may identify more diverse immune cell populations, and that the logistics of freezing and thawing may impact composition, especially for granulocyte populations. Finally, we were able to identify a distinct subset of macrophages defined by elevated co-expression of multiple inflammatory cytokines, including *IL1B, CCL3, CCL20, CCL3L1*, termed “Cytokine Expressing Macrophages”.

**Figure 1.**
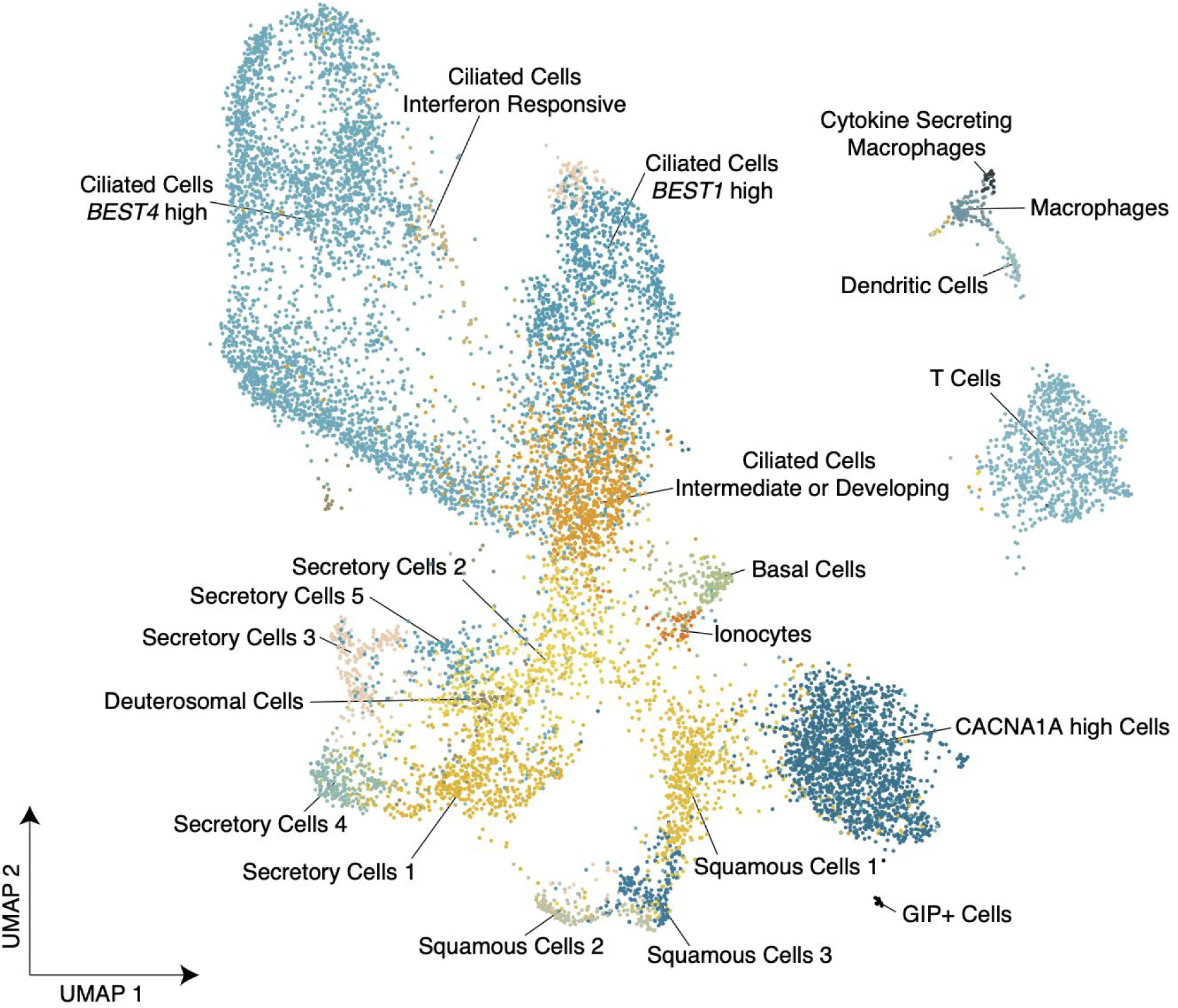
A single-cell representation of nasopharyngeal swabs from control and COVID-19 participants sampled at the University of Mississippi Medical Center. Uniform manifold approximation and projection (UMAP) of 12,427 single cells, with points colored by cell identity (**Supplementary Table 1**), representing n= 6 non-COVID-19 SARS-CoV-2-negative controls, and n= 15 COVID-19 participants PCR-positive for SARS-CoV-2.

Among epithelial cell clusters, we recovered ciliated cells, secretory/goblet cells, squamous cells, and basal cells. We also recovered deuterosomal cells, defined by expression of *DEUP1, CDC20B, HELLS, FOXN4*, and *HES6*, that were recently identified by Garcia et al. using scRNA-seq of airway epithelial cultures (*Garcia et al., 2019*). This population is thought to represent an important precursor population for ciliated cell types through upregulation of genes required for centriole amplification and cilium assembly. Ionocytes were also readily distinguished from other epithelial cell types by expression of *FOXI1, FOXI2, CFTR*, and *ASCL3*. Ciliated epithelial cells (defined by unique expression of *PIFO, CAPS, FOXJ1*, and multiple gene modules encoding components of cilia microtubule structures) were the most abundant cell type recovered across all nasal swabs. Upon subclustering of ciliated epithelial cells, we observed four distinct cell subtypes, each defined by significant expression of unique genes, and putatively named “Ciliated Cells *BEST4* high”, “Ciliated Cells *BEST1* high”, “Ciliated Cells Intermediate or Developing”, and “Ciliated Cells Interferon Responsive”. At present, we are unsure as to whether the “Ciliated Cells Intermediate or Developing” represent a developmental precursor to more terminally differentiated ciliated cell types, or are a technical artifact of cellular complexity or gene spillover, or a combination of both. Further work is necessary to model developmental trajectories within this dataset.

Squamous cell clusters were annotated by their unique expression of genes such as *SCEL, KLK7, KRT6A*, and subdivided into 3 clusters, temporarily named “Squamous Cells 1”, “Squamous Cells 2”, and “Squamous Cells 3”. In general, cells within “Squamous Cells 1” appear to express diminished abundances of genes present in more differentiated squamous cell clusters. As discussed with respect to “Ciliated Cells Intermediate or Developing” above, further analysis and additional samples will be required to resolve whether this represents a developmental intermediate or technical artifact. Notable defining genes expressed by “Squamous Cells 2” include cytokines *IL36G, IL36A*, and *CXCL1*, as well as highest expression of cornified envelope genes *SPRR2D, SPRR1B, SPRR2A, SPRR3, SPRR2E* and *CNFN*, potentially indicating this subset of cells is at a later stage in squamous cell development. “Squamous Cells 3” express higher levels of interferon response genes than other squamous cell clusters, including *OAS3, OAS1*, and *MX1*, and multiple genes involved in antigen presentation and processing, such as *TAPBP, TAP1, HLA-C*, and *HLA-F*, as well as *VEGFA*.

We identified substantial diversity among Secretory/Goblet cell populations within this dataset, putatively named “Secretory Cells 1-5”, which all express high abundances of *AQP5, CXCL17, SLPI*, and *PIGR*. “Secretory Cells 1” and “Secretory Cells 2” express high levels of *MUC16, MUC4 and MUC5AC* compared to other secretory populations (many of which express non-zero UMI of these mucins, but at diminished frequency and abundance). “Secretory Cells 2” are distinguished by high expression of various interferon responsive genes such as *IFI44, STAT6, DUOX2, HLA-F, HLA-DRA, HLA-A*, and *HLA-DRB5*, as well as *TNFSF10* (TRAIL). Our current dataset suggests “Secretory Cells 1” and “Secretory Cells 2” represent two types of goblet cells, and based on expression of *SCGB1A1* within “Secretory Cells 2” and “Secretory Cells 3”, there are potentially club cells intermixed within these clusters (or these clusters represent developmental intermediates), which may be resolved with additional samples and deeper profiling. Like “Secretory Cells 1” and “Secretory Cells 2”, “Secretory Cells 3” also express high abundances of *BPIFB1*, but lack high expression levels of other mucins. “Secretory Cells 3” also express high levels of antigen processing and presentation related genes and interferon response factors, and uniquely contain high levels of early response genes and AP-1 family transcription factors such as *FOSB, MAFF, DUSP1, KLF4, NR4A1, ATF3, JUND, ID3*, and *ID1*. “Secretory Cells 4” are distinguished by expression of *PLAU* and *SAA1*/*SAA2*, two acute phase response proteins, in addition to *NFKBIA, NFKBIZ, CXCL8, CXCL2* and *IL32*. Finally, “Secretory Cells 5” are unique from other secretory cells by their elevated expression of serine protease inhibitors *SERPINB13, SERPINB4, SERPINB3* among other genes and pathways.

Finally, we recovered two unexpected cell types within this dataset, temporarily named “*CACNA1A* high Cells” and “*GIP+* Cells”. Both are defined by expression of *CACNA1A*, and *GIP*+ cells express *GIP* and *LGR5*.

#### Analysis of SARS-CoV-2 Viral Transcripts and Host Cells

We found that cells containing UMI aligning to SARS-CoV-2 genomic features were restricted to swabs from individuals diagnosed with COVID-19, and were not detected in any cells from control nasal swabs. Among 15 study participants in the current dataset diagnosed with COVID-19, nasal swabs from 9 contain high quality single-cell transcriptomes with reads aligning to SARS-CoV-2. Notably, all participants diagnosed with COVID-19 had received a positive SARS-CoV-2 PCR from nasopharyngeal swab within 3 days of sample collection for this study. Among the 6 nasal swabs from study participants with COVID-19 where we did not detect SARS-CoV-2 viral RNA+ cells, 3 had poor recovery of high-quality single cell transcriptomes, diminishing our power to detect rare viral reads if present.

Among swabs from the COVID-19 group containing any viral RNA+ cell, the percentage of infected cells across all cell types was 12.9 +/- 7.1 % (mean +/- SEM, n=9). This corresponded to a number of SARS-CoV-2 RNA+ cells ranging from 1 to 126 per nasal swab, with an average +/- SEM of 30.2 +/- 13.0 cells. SARS-CoV-2 viral aligning UMI were found in multiple cell types. The highest proportions of viral RNA+ cells per cluster were found in “Secretory Cells 5” (23.8% of cluster, across all COVID19 samples), “Ciliated Cells Interferon Responsive” (20.8%), “Secretory Cells 1” (8.7%) and “Cytokine Secreting Macrophages” (7.4%), and, to a lesser extent, “Ciliated Cells Intermediate or Developing” (6.4%), “Macrophages” (6.3%), and “Basal Cells” (4.3%). Within each of these clusters, we identified cells containing a high total abundance of viral reads and viral RNA+ single cells consistently contained UMI aligning to diverse regions across the viral genome, increasing our confidence for true viral genomic capture from intact virions or viral RNA completing a replication cycle, rather than spurious viral RNA derived from ambient pools of RNA. “Ciliated Cells BEST1 high” and “Ciliated Cells BEST4 high” additionally contained high absolute abundances of SARS-CoV-2 RNA+ cells, however the within-cluster proportion of these cells was low (<5%).

### 4.4 University College London & Wellcome Sanger Institute

#### Study Overview and Design

Subjects 18 years and older were included from two large hospital sites in London, United Kingdom, namely University College London Hospitals NHS Foundation Trust and Royal Free London NHS Foundation Trust during the height of the pandemic in the United Kingdom (April to July 2020). Ethical approval was given through the Living Airway Biobank, administered through UCL Great Ormond Street Institute of Child Health (REC reference: 19/NW/0171, IRAS project ID 261511), as well as by the local R&D departments at both hospitals. At daily virtual COVID19 coordination meetings, suitable patients were chosen from a list of newly diagnosed and admitted patients within the preceding 24 hours (based on a positive nasopharyngeal swab for SARS-CoV-2). Patients with typical clinical and radiological COVID19 features but with a negative screening test for SARS-CoV-2 were excluded. Other excluding criteria included active haematological malignancy or cancer, known immunodeficiencies, sepsis from any cause and blood transfusion within 4 weeks. Maximal severity of COVID19 was determined retrospectively by determining the presence of symptoms, the need of oxygen supplementation and the level of respiratory support (reference joint table of severity). Nasal brushings and peripheral blood sampling were performed by trained clinicians prior to inclusion to any pharmacological interventional trials within 48 hours of positive SARS-CoV-2 nasopharyngeal sampling.

#### Initial Observations

The first release of data includes samples from 12 adults, 12 blood with 3 matched nasal samples. The cohort comprised 5 males and 7 females, aged between 25 and 76 years old. Disease severity varied from mild to moderate and severe disease, with severity coded according to WHO standards. Analysis of both the blood and nasal samples is ongoing and cell type annotation at this stage is very preliminary. More fine-grained annotations will be provided in future data releases. For the nasal samples a range of epithelial and immune cells were identified. The immune cell population included B cell, T cells, NK cells, dendritic cells, macrophages, monocytes, dendritic cells, mast cells and neutrophils, all identified by previously described marker genes. Using nasal brushes, we also retrieved a variety of epithelial cells, which included basal cells, cycling basal cells, squamous cells and three populations of secretory cells. Two populations of ciliated cells could be distinguished, as well as deuterosomal cells, likely intermediates between secretory and ciliated cells, and ionocytes.

### 4.5 VIB & Ghent University Hospital

#### Study Overview and Design

We profiled matching bronchoalveolar lavage fluid (BALF) and blood samples from patients who have been hospitalized with a high clinical suspicion of COVID-19 (n=17) and control individuals (n=2). The analysis includes single-cell 3’ RNA-sequencing along with the quantitative measurement of surface proteins using panels of more than 250 oligo-conjugated antibodies (TotalSeq A - CITEseq). The study population entails adult patients with a diagnostic or therapeutic need for bronchoscopy and the cohort consists of COVID-19 patients (n=8), control cases with a non-SARS-CoV-2 respiratory disease (n=9) and healthy controls (n=2). Patients aged 18-100 years old were eligible for study inclusion if they had clinical symptoms suggestive of COVID19 and if hospitalization was required. Healthy controls were asymptomatic and were selected from a group of patients requiring a bronchoscopy with BAL for diagnostic work-up or follow-up of other diseases. In these cases, lavage was always performed in a healthy lung lobe and SARS-CoV-2 was formally ruled-out by rRT-PCR. For 5 out of 19 patients, we analyzed PBMC samples at a secondary time point as well. This study was performed in accordance with the principles expressed in the Declaration of Helsinki. Written informed consent was obtained from all patients or a legal representative. The study was approved by the Ethics Committee of Ghent University Hospital (Belgium), AZ Jan Palfijn (Belgium) and AZ Maria Middelares (Belgium), where all samples have been collected.

#### Initial Observations and Challenges

The first interim dataset of BALF samples consists of 275.056 cells derived from 19 study participants. The cohort consists of COVID-19 patients (n=8), control cases with a non-SARS-CoV-2 respiratory disease (n=9) and healthy controls (n=2). The current data release contains a first cell type annotation based on the gene/antibody expression profiles. This annotation requires additional verification, both in terms of cell quality and cell identity. Efforts are underway for updated annotations after further iterations of this dataset, and inclusion of patient-matched PBMC samples (sampled at the time of the bronchoalveolar lavage and at later time point, totalling 377.684 cells derived from the blood). Updates and additional clinical metadata are available through the portals www.single-cell.be/covid19 and www.covid19cellatlas.org. Comparison between COVID19 positive and COVID19 negative patients shows a distinct bias towards more severe disease status, longer time since onset of symptoms and longer time since hospital admission in the COVID19 positive group. Furthermore, at the time of collection of BALF samples 75% (6/8) of the COVID19 positive patients were mechanically ventilated and admitted into an intensive care unit (ICU), whereas none of the COVID19 patients were admitted into ICU. Therefore, we are currently expanding this cohort of BALF samples to patients in the ICU with ARDS based on non-COVID-19 pathology, to better control for this bias. After expanding this dataset, we will perform in-depth compositional analyses across COVID-19 samples and disease groups.

After dimensionality reduction, integration and clustering of the BALF cells, we mapped more than 60 clusters corresponding with immune and epithelial cell identities in our preliminary analysis defined by expression of specific marker genes or antibodies. Epithelial cell types include AT1 (*AGER, HOPX*) and AT2 (*NAPSA, SFTB*) cells, ciliated cells (*FOXJ1, CAPS*), basal cells (*KRT5, DAPL1*), ionocytes (*FOXI1, ASCL3*), mucous (*MUC5AC, SCGB3A1*) and serous (*PRR4, LYZ*) secretory cells, deuterosomal cells (*DEUP1, PLK4*). We recovered a CD309high *SPRR3*high fraction of epithelial cells reminiscent to previously described mouse “hillock” cells (*Deprez et al., 2020*). Three granulocyte subtypes were identified: neutrophils (based on CD16, CD66b and CD15 protein levels), basophils (*CPA3*, CD123) and mast cells (*CPA3*, CD117). A preliminary phenotyping of CD4 T cell subsets revealed naive (*CCR7*), exhausted (*LAG3, CTLA4*), activated (*TNFRSF9*), cytotoxic (*GNLY*), regulatory (*FOXP3, CTLA4*) and follicular helper-like T cells (*CXCL13, IL21, CD278*). Other lymphoid identities include activated, naive (*CCR7*) and EMRA (*FGFBP2*) CD8 T cells; MAIT cells (*KLRB1, TRDC*); various subtypes of NK/iNKT cells; B cells (*CD19, CD79A*), and plasmablasts/plasma cells (*MRB1*). The dendritic cell types include cDC1 (*CLEC9A*), cDC2A (*RUNX3, SREBF2*), cDC2B (*CD1C, FCER1A, CD5*), cDC3 (*CD14, CD163, CLEC12A*), migratory cDCs (*CCR7*) and pDCs (*MZB1, IRF8*). Additionally, we recovered mature (*C1QA*), resident (*MARCO*) and alveolar (*FABP4*) macrophages next to recruited activated (*VCAN, CCL2*) and patrolling (*HSPA6*) monocytes. Thus far, we did not detect reads aligning to the SARS-CoV-2 transcriptome in these or any cells in this data set.

### 4.6 Massachusetts General Hospital & Broad Institute

#### Study Overview and Design

The vast majority of deaths are due to acute lung injury and acute respiratory distress disorder (ARDS), or direct complications thereof (*Zhou F et al., 2020; Wölfel R et al. 2020; Xu Z et al. 2020; Yang X et al.2020; Yang F et al. 2020; Du et al. 2020*). The progression to ARDS is thought to reflect a combination of increasing viral load, cytopathic effects, translocation of virus into pulmonary tissue, including infection of pulmonary endothelial cells, and inappropriate or insufficient immune responses. Since ARDS is an acute inflammatory injury to the lung, we hypothesize that COVID-19 associated ARDS develops, at least in part, as a direct result of a dysfunctional host immune response that contributes to clinical deterioration in the acute phase of systemic illness, leading to ineffective viral clearance and collateral pulmonary or other tissue damage. Understanding immune signaling that is enhanced or suppressed in patients with worse outcomes has the potential to identify new therapeutic targets for COVID-19 associated ARDS, and knowing the kinetics of potentially pathogenic immune responses will inform the optimal timing of interventions.To address this, the immune and cellular response of infected patients needs to be analyzed comprehensively. Single-cell multi-omics strategies are uniquely poised to do this, given the substantial cellular complexity and the minute clinical samples. Our goals are to rapidly define immune cell states and signaling pathways in COVID-19 that are associated with ARDS severity and to investigate the role of these pathways in ARDS. To achieve this, we enrolled patients in the Emergency Department (ED) in a large, urban, academic hospital from 3/24/2020 to 4/30/2020 in Boston during the peak of the COVID-19 surge, with an institutional IRB-approved waiver of informed consent. We included patients 18 years or older with a clinical concern for COVID-19 upon ED arrival, and with acute respiratory distress with at least one of the following: 1) tachypnea ≥ 22 breaths per minute, 2) oxygen saturation ≤ 92% on room air, 3) a requirement for supplemental oxygen, or 4) positive-pressure ventilation. A blood sample was obtained in a 10 mL EDTA tube concurrent with the initial clinical blood draw in the ED. We enrolled 384 unique subjects who presented to our ED with acute respiratory distress suspected or known to be due to COVID-19. 306 patients were subsequently confirmed to have COVID-19 infection. In addition to blood samples collected in the ED (D0, n=384 samples), blood draws were also obtained at Day 3 (n=220 samples) and Day 7 (n=141 samples) of hospitalization, as well as at event-driven timepoint (i.e. decompensation or recovery events) (n=43 samples) obtained anywhere within the first 28 days of enrollment. For every blood sample collected, we isolated and cryopreserved peripheral blood mononuclear cells (PBMCs) for downstream single-cell RNA-sequencing (*Zheng et al. 2017*) with paired CITE-seq (*Stoeckius et al. 2017; Peterson et al. 2017*), TCR and BCR measurements. We also cryopreserved plasma to perform proteomics analyses, for which data is already available for download ([MGH COVID-19 study proteomics data](https://info.olink.com/mgh-covid-study-overview-page?hsCtaTracking=fff99a2a-81c1-4e4a-a70d-6922d26503b4%7C202c2809-0976-48f7-aad0-3903c36624ca)).

Clinical course for every patient was followed to 28 days post-enrollment, or until hospital discharge if that occurred after 28 days. Of all 384 enrolled, 78 (20%) tested negative for SARS-CoV-2 during their hospitalization. Of the patients who tested negative for SARS-CoV-2, 50 (64%) had very low suspicion for COVID-19 based on careful retrospective chart review. These 50 subjects were categorized as controls, for which single-cell RNA sequencing data is being generated. For the remaining 28 SARS-CoV-2 negative patients, COVID-19 was a diagnostic possibility yet most had multiple negative PCR tests throughout their hospital course. We classified SARS-CoV-2 positive subjects by illness severity and outcome derived from the World Health Organization (WHO) Ordinal Outcomes Scale (WHO R&D Blueprint - Novel Coronavirus, COVID-19 Therapeutic Trial Synopsis.), which can be on a 6-point scale. We classified patients by acuity levels on days 0, 3, 7, and 28, derived from the WHO Ordinal Outcomes Scale. Our primary outcome for confirmed COVID-19 cases was the maximal acuity within 28 days of enrollment: WHO-1, death within 28 days (N=42, 14%); WHO-2, intubation, mechanical ventilation, and survival to 28 days (N=67, 22%); WHO-4, hospitalized and requiring supplemental oxygen (N=133, 43%); WHO-5, hospitalized without requiring supplemental oxygen (N=41, 13%); and WHO-6, discharged directly from the ED without returning within 28 days (N=23, 8%). We note that we did not collect any samples from WHO category 3, consisting of patients who would have been hospitalized and requiring non-invasive ventilation. Demographic, past medical history and clinical data were collected and summarized for each outcome group, using medians with interquartile ranges where appropriate. The meta-data accompanying the dataset is described in the table below.

#### Initial Observations and Words of Caution

The first released dataset includes data from 32 PBMCs collected from 15 patients across multiple time points. This single-cell dataset includes 59,506 cells after excluding cells of lower quality. **Importantly, the current release contains a preliminary cell type annotation on the cluster level based only on the gene expression profiles**. This annotation still requires additional verification, both in terms of cell quality and cell identity, as well as properly identifying potential doublets. Future data releases will include additional data as well as an annotation based on expression profiles of both gene and cell surface markers (i.e. we measured 197 surface proteins through CITE-seq) in relevant subsets of the data. Our preliminary analysis of peripheral blood captured all known populations. The myeloid cell fraction is comprised of conventional dendritic cells (“cDC”: *CD1C, FCGR2B, CD1E*), plasmacytoid dendritic cells (“pDC”: *IL3RA, CLEC4C, TCF4*), as well as 5 putative monocyte-like populations: “Monocyte_1” (*CD14, LYZ, ASGR1*), “Monocyte_2” (*S100A8, S100A9, CD163, IFITM3*), “Monocyte_3” (*FCGR3A*), “Monocyte_4” (*CD163, IER3*), “Monocyte_5” (*C1QA, C1QB, C1QC*). These myeloid cell populations were notably diverse compared to what has been observed in blood analyzed from “healthy” individuals. Efforts are underway to provide a more comprehensive annotation of myeloid subsets in this cohort through additional data generation and subclustering. The T cell fraction captures CD4+ T cell subsets – including for example naive (*CCR7*) and Treg (*FOXP3, IL2RA*) CD4+ T cells –, CD8+ T cell subsets, as well as cycling CD8 T cells (“CD8_T cell 2”; *MKI67, STMN1*). B cell initial subsets observed include naive B cells (“B cell_2”: *TCL1A, FCER2*), memory B cells (“B cell_1”: *CD27, CD80, LSP1*); and plasma cells (*JCHAIN, MZB1*). Additionally, we identified NK cells (*NCAM1, FCGR3A*) and platelets (*PPBP, PF4*). Although not included in our original data release, efforts are underway to provide a more comprehensive annotation of immune cell subsets in these specimens, including through sub-clustering analyses. Furthermore, we suspect further iterations of this dataset with the larger sample size being generated from the entire cohort may identify more diverse immune cell populations.

## 5. Materials and Methods

### UMMC/BCH/Broad

### Next Steps

We are currently expanding this cohort of nasal swabs from 21 participants to at least 40, across both COVID-19 and Control groups. Where possible, we will also be adding endotracheal suction samples. After generating data from this expanded cohort, we will revise our cellular clusters and perform deeper, more powered analyses into differences in cellular composition across disease groups and within samples from COVID-19 participants. We will comprehensively analyze SARS-CoV-2 RNA+ cells and identify cell-type-specific host programs associated with viral genomic material.

### Summary of Methods

#### Words of Caution

Nasopharyngeal Swabs: These data represent single cells recovered by nasopharyngeal swab, and it is important to remember that cell recovery and composition will vary compared to nasal scrapings (which typically access the turbinates) or nasal washes, and may be influenced by donor specific factors (e.g., age, time since infection, comorbidities, etc). Additionally, this release represents approximately half of the samples to be included in the complete dataset, and changes in the diversity of cell types – particularly detection of rare cell subsets – are expected following creation of a larger dataset. Finally, preliminary analyses of SARS-CoV-2 aligning UMIs identify select cellular clusters thought to harbor the majority of virally targeted cell types or cell types which have engulfed virus or infected cells. We are in the process of applying a more robust and conservative statistical procedure to correct for possible ambient RNA contamination, which will be critical for downstream analyses of the association between SARS-CoV-2 viral RNA and host biology.

### Nasal Swabs

#### Sample collection and tissue dissociation (Sanger/Barcelona)

Detailed sample processing for blood samples and nasal swabs are available through protocols.io (dx.doi.org/10.17504/protocols.io.bjinkkde). Briefly, PBMCs were obtained from peripheral blood by Ficoll gradient using Lymphocyte Isolation Solution. Once PBMCs were isolated, all samples were stored at −150°C in fetal bovine serum (FBS) + 10% DMSO. Cryopreserved PBMCs were thawed rapidly in a 37°C water bath and then slowly diluted in pre-warmed growth medium, centrifuged and resuspended in fresh FACS buffer (PBS + 3% FBS) prior to loading them into the 10X Chromium. Additionally, a fraction of the thawed PBMCs was depleted for CD3+ cells, using anti-CD3+ magnetic beads prior to loading into 10X Chromium.

Nasal swabs were transported in medium (MEM + 1% penicillin/streptomycin + 0.1% amphotericin + 0.1% gentamicin) from the hospital to the laboratory. Cell clamps were then digested with B. licheniformis protease (10 mg/mL) on ice for 30 minutes. After quenching the protease, single cell suspensions were treated to remove red cells using eBioscience 1x red blood cell (RBC) lysis buffer (Invitrogen) and filtered using a Flowmi strainer. Finally, cells were counted to achieve the desired cell concentration.

### UMMC/BCH/Broad Team (Nasopharyngeal Swabs Methods)

#### Sample Collection and Biobanking

Nasopharyngeal samples were collected by trained healthcare providers using FLOQSwabs (Copan flocked swabs) following the manufacturer’s instructions. Collectors would don personal protective equipment (PPE), including a gown, non-sterile gloves, a protective N95 mask, a bouffant, and a face shield. The patient’s head was then tilted back slightly, and the swab inserted along the nasal septum, above the floor of the nasal passage to the nasopharynx until slight resistance was felt. The swab was then left in place for several seconds to absorb secretions and slowly removed while rotating swab. A second swab was then completed in the other nares. The swabs were then placed into a cryogenic vial with 900 µL of heat inactivated fetal bovine serum (FBS) and 100 µL of dimethyl sulfoxide (DMSO). The vials were then placed into a Thermo ScientificTM Mr. Frosty Freezing Container for optimal cell preservation. The Mr. Frosty containing the vials was then placed in cooler with dry ice for transportation from patient area to laboratory for processing. Once in the laboratory, the Mr. Frosty was placed into the −80°C Freezer overnight and then on the next day, the vials were moved to the liquid nitrogen storage container.

#### Dissociation and Collection of Viable Single Cells from Nasal Swabs

Swabs in freezing media (90% FBS/10% DMSO) were stored in liquid nitrogen until immediately prior to dissociation. A detailed sample protocol can be found here: dx.doi.org/10.17504/protocols.io.bjhmkj46. This approach ensures that all cells and cellular material from the nasal swab (whether directly attached to the nasal swab, or released during the washing and digestion process), are exposed first to DTT, followed by an Accutase digestion. Briefly, nasal swabs in freezing media were thawed, and each swab was rinsed in RPMI before incubation in 1 mL RPMI/10 mM DTT (Sigma) for 15 minutes at 37ºC with agitation. Next, the nasal swab was incubated in 1 mL Accutase (Sigma) for 30 minutes at 37ºC with agitation. The 1 mL RPMI/10 mM DTT from the nasal swab incubation was centrifuged at 400 g for 5 minutes at 4ºC to pellet cells, the supernatant was discarded, and the cell pellet was resuspended in 1 mL Accutase and incubated for 30 minutes at 37ºC with agitation. The original cryovial containing the freezing media and the original swab washings were combined and centrifuged at 400 g for 5 minutes at 4ºC. The cell pellet was then resuspended in RPMI/10 mM DTT, and incubated for 15 minutes at 37ºC with agitation, centrifuged as above, the supernatant was aspirated, and the cell pellet was resuspended in 1 mL Accutase, and incubated for 30 minutes at 37ºC with agitation. All cells were combined following Accutase digestion and filtered using a 70 µm nylon strainer. The filter and swab were washed with RPMI/10% FBS/4 mM EDTA, and all washings combined. Dissociated, filtered cells were centrifuged at 400 g for 10 minutes at 4ºC, and resuspended in 200 µL RPMI/10% FBS for counting. Cellular recovery, estimated viability percentages, and representative images are being compiled and will be shared with final datasets. Cells were diluted to 20,000 cells in 200 µL for scRNA-seq. For the majority of swabs, fewer than 20,000 cells total were recovered. In these instances, all cells were input into scRNA-seq.

### UCL/Sanger

Samples were collected and transferred to a Category Level 3 facility at University College London and processed within 2 hours of sample collection. Nasal brushings were enzymatically digested to a single cell solution and processed further straight away. Peripheral blood was centrifuged after adding Ficoll Paque Plus and PBMCs, serum and neutrophils separated, collected and frozen for later processing.

### BALF Protocols

#### Sample Preparation (Columbia)

Detailed sample processing protocols for both airway washes and blood samples have been deposited on protocols.io (airway: dx.doi.org/10.17504/protocols.io.bjj8kkrw; **blood:** dx.doi.org/10.17504/protocols.io.bjm6kk9e). Briefly, for the airway washes, we treated samples with Benzonase, filtered, and then isolated mononuclear cells (MNCs) with a Ficoll-Paque PLUS density gradient. To remove neutrophils and red blood cells from the MNC preparation, we treated with biotinylated anti-CD66b and anti-CD235ab, respectively, and depleted antibody-bound cells with streptavidin-coated magnetic beads. We also depleted the MNCs of dead cells using Dead Cell Removal Microbeads (Miltenyi). For the blood samples, we isolated MNCs using a Ficoll-Paque PLUS density gradient, but included the RosetteSep Granulocyte Depletion Cocktail (Stemcell Technologies) to remove neutrophils. We further purified the MNCs using anti-CD66b/anti-CD235ab magnetic depletion and dead cell removal as described above for the airway washes.

#### Single-Cell Capture Method

We used a Next GEM Chromium Controller (10x Genomics) to capture and lyse individual cells and for co-encapsulation with barcoded primers. We loaded the instrument to target ∼5,000 scRNA-seq profiles per lane after cell counting with NucleoCounter NC-3000 (ChemoMetec).

### PBMC

#### Sanger/IJC Group

PBMCs and CD3+-depleted PBMCs (CD3N) (**dx.doi.org/10.17504/protocols.io.bjinkkde**) were stained with CITEseq antibodies and loaded into the 10X Chromium machine. In some cases, PBMCs or CD3N cells from two different donors were multiplexed under the same reaction. Detailed sample staining is available through protocols.io. Briefly, for the CITEseq protocol (**dx.doi.org/10.17504/protocols.io.bjhmkj46**), cells were resuspended in FACS buffer (PBS + 4% FBS), incubated with Fc Block for 10 minutes and the specific mix of antibodies for 30 minutes at 4ºC. Cells were then washed three times, filtered using a Flowmi strainer and counted.

Cells were loaded into the instrument to target ∼25,000 scRNA-seq profiles per lane. In the case of nasal swabs, fresh cells were loaded into the 10X Chromium machine to target between 1,000 and 5,000 scRNA-seq profiles.

### Library Preparation

#### Library Generation and Sequencing (Columbia & Sanger)

We used the Chromium Next GEM Single Cell 3’ Reagent Kit v3.1 (10x Genomics) for scRNA-seq library construction and sequenced the resulting libraries on an Illumina NovaSeq 6000. We targeted ∼300M raw reads per sample (∼60,000 raw reads per cell) with cycle numbers of 100 for read 1, 100 for read 2, and 100 for the index read.

### Blood sample collection and PBMC isolation *(MGH/Broad Acute Blood Cohort)*

Blood samples were collected in EDTA tubes, and processed no more than 3 hours post blood draw in a Biosafety Level 2+ laboratory on site. Peripheral blood mononuclear cells (PBMC) were isolated from whole blood using Ficoll gradient centrifugation protocol detailed in protocol.io: dx.doi.org/10.17504/protocols.io.bjhnkj5e. Briefly, whole blood was diluted with room temperature RPMI medium in a 1:2 ratio to facilitate cell separation for other analyses using the SepMate PBMC isolation tubes (STEMCELL) containing 16ml of Ficoll (GE Healthcare). Diluted whole blood was centrifuged at 1200 rcf for 20 minutes at 20°C. After centrifugation and additional washes detailed in the protocol, aliquots of plasma and PBMCs were prepared and cryopreserved for downstream analyses. The PBMC aliquots were placed into a Thermo ScientificTM Mr. Frosty Freezing Container for optimal cell preservation overnight into the −80°C Freezer. The vials were moved to the liquid nitrogen storage container. As detailed in our protocol in protocol.io: **dx.doi.org/10.17504/protocols.io.bjhnkj5e**, neutrophils were also isolated from every whole blood sample and lysed for downstream bulk-RNA sequencing analysis.

### PBMCs processing for single-cell genomics analysis *(MGH/Broad Acute Blood Cohort)*

We processed every PBMC samples for single-cell RNA sequencing (scRNAseq) with paired measurements of 197 surface proteins (CITE-seq, using TotalSeq reagents from Biolegend), T cell receptor (TCR) and B cell receptor (BCR) sequencing using the 10X Genomics Next GEM Single Cell 5’ Reagent Kit v1.1 (see below for further details). PBMC samples were processed by a batch of 8 samples. For every batch, several clinical meta-data were taken into account to limit batch effect, including balancing for: gender, age range, WHO groups, and pre-existing conditions. For each batch of 8 samples, upon thawing up to 1.5M cells per sample and performing an initial washing step through centrifugation at 300g for 7min at 4°C, dead cells (EasySep Dead Cell Removal Kit; Stemcell, catalog #17899) and red blood cells (ErythroClear Read Blood Cell Depletion Kit; Stemcell, catalog #01738) were depleted from each sample using commercially available kits. The total number of remaining viable cells were counted using trypan blue. Up to 250,000 cells per sample were then selected to be stained with CD45 selection mastermix (MojoSort CD45 Nanobeads, BioLegend, catalog #480030; Human TruStain FcX, Biolegend, catalog #422302) together with TotalSeq-C Hashtags (BioLegend). MojoSort CD45 Nanobeads were used to help capture cells over debris for downstream analyses. Eight distinct TotalSeq-C hashtags were selected for generating data (i.e. TotalSeq-C0251, C0252, C0254, C0256, C0257, C0258, C0259, C0260). After performing three washes, cells were counted with trypan blue. 60,000 cells from each of the 8 samples were then pooled together to a 1.5 ml lo-bind eppendorf tube. Upon filtering the pooled cell suspension with a 40 um Bel-art Flowmi strainer into a new 1.5 ml lo-bind eppendorf tube, the cell suspension was stained with a customized TotalSeq-C 197-antibody pool (Biolegend), following the manufacturer’s instructions for concentration. Pooled cells were incubated on ice for 30 minutes; cells were resuspended every 10 minutes. Four washing steps were then performed. After completing the last washing steps, the cell pellet was resuspended in 50uL of RPMI-1640 medium (Thermo Fisher Scientific) supplemented with 2% human AB serum (Sigma-Aldrich). Cells were counted and concentration adjusted to load 50,000 cells on the 10X Genomics Chromium Controller (see below).

### Library Generation and Sequencing (MGH/Broad Acute Blood Cohort)

A Next GEM Chromium Controller (10x Genomics) was used to capture single cells with a target of 3,000 scRNAseq profile per PBMCsample. We used the Chromium Next GEM Single Cell 5’ Reagent Kit v1.1 (10x Genomics) for scRNA-seq library construction, according to the manufacturer’s instructions. Libraries were multiplexed using individual Chromium i7 Sample Indices, and then sequenced on NovaSeq S4 Flowcells with custom sequencing metrics (single-indexed sequencing run, 28/8/0/96 cycles for R1/i7/i5/R2), aiming for 50,000 reads per cell for gene expression, 7000 reads per cell for TCR, 7000 reads per cell for BCR, and 10,000 reads for the feature barcodes (FBC).

### Sample Collection and Processing for CITEseq/scRNAseq (BALF cohort - VIB / Ghent University Hospital)

Bronchoscopy with BAL was performed bedside using a single use disposable video bronchoscope. Bronchoscopy was only performed in hemodynamically and respiratory stable patients. In spontaneously breathing patients, an additional oxygen need of 3L/min in rest was required. Recommended personal protective equipment was used: full face mask, disposable surgical cap, medical protective mask (N95/FFP2/FFP3), work uniform, disposable medical protective gown, disposable gloves. Three to five aliquots of 20 mL sterile normal saline were instilled into the region of the lung with most aberrations on chest CT. Retrieval was done by suctioning of the scope. BAL fluid was collected in siliconized bottles to prevent cell adherence and kept at 4 °C. BAL fluid was filtered through a 100 μm cell strainer (BD Biosciences) and centrifuged for 7 min at 1300 rpm at 4 °C. The supernatant was removed and the BAL fluid cells were counted and subsequently processed fresh for CITEseq/scRNAseq. One million of cells was used for subsequent single cell RNA sequencing while the remaining cells were frozen in 1 mL 90% fetal calf serum (FCS, Sigma), 10% dimethyl sulphoxide Hybri-Max (DMSO, Sigma) in a cryovial using a 5100 Cryo 1 °C Freezing Container (Nalgene) to - 80 °C. Afterwards the cells were stored stored in liquid nitrogen (196°C). Whole blood was collected in EDTA tube and processed within a maximum of 1.5 hours after collection. Whole blood separation was performed by bringing whole blood, diluted with PBS 7.2 (ThermoFisher Scientific, # 20012027), in a Leucosep™ tube, (Greiner Bio-One, # 227290), prefilled with 15 mL Lymphoprep™(Stemcell technologies, # 07851), followed by a centrifugation step of 30 minutes at 1500 rpm (acceleration 5, brake 3). After isolation, the PBMCs were twice washed in PBS 7.2 and centrifuged at 350 xg for 10 minutes in a cooled centrifuge at 4°C. Isolated PBMCs were counted, cryopreserved in 1mL FCS/DMSO 10% and stored in liquid nitrogen (196°C).

### Single-Cell Capture Method and Library Preparation (BALF cohort - VIB / Ghent University Hospital)

All experiments have been conducted at a containment laboratory with inward directional airflow (BSL-3). BALF cells have been processed fresh, PBMC cells were frozen first and subsequently processed. One million of cells was stained with the CITE-seq antibody mix containing >250 barcoded antibodies (TotalSeq™-A, BioLegend), CD45 FITC (Clone HI30, BioLegend, 3040050), and CD235a APC (2.5uL, Clone HIR2, BD Biosciences, 561775). When cell hashing was applied, TotalSeq™-A hashing antibodies were supplemented to the CITEseq antibody cocktail. After a 30 min incubation on ice, cells were then washed with PBS/FBS2% and spun down at 500 rcf at 4°C for 5min. After resuspension in 300uL of PBS and instant staining with propidium iodide (Company, catalog number, 4uL), PI-/CD235a-viable cells (whilst excluding red blood cells) were sorted using the BD FACSJazz™. Sorted cells were spun down at 450rcf at 4°C for 8min. Supernatant was carefully discarded and the cell pellet was resuspended in an appropriate volume of PBS/BSA 0.04%. Sorted cells were loaded on a GemCode NextGEM Single-Cell Instrument (10x Genomics) to generate single-cell Gel Bead-in-EMulsion (GEMs). and samples were mixed prior loading on the GemCode instrument. Single-cell RNA-Seq libraries were prepared using GemCode Single-Cell V3.1 (NextGEM) 3’ Gel Bead and Library Kit (10x Genomics) according to the manufacturer’s instructions. Sequencing libraries were sequenced with NovaSEQ S4 flow cell with custom sequencing metrics (single-indexed sequencing run, 28/8/0/98 cycles for R1/i7/i5/R2) (Illumina). Sequencing was performed at the VIB Nucleomics Core (VIB, Leuven, Belgium).

### scRNA-seq (UMMC/BCH/Broad)

Seq-Well S3 was run as previously described. Briefly, a maximum of 20,000 single cells were deposited onto Seq-Well arrays preloaded with a single barcoded mRNA capture bead per well. Cells were allowed to settle by gravity into wells for 10 minutes, after which the arrays were washed with PBS and RPMI, and sealed with a semi-permeable membrane for 30 minutes, and incubated in lysis buffer (5 M guanidinium thiocyanate/1 mM EDTA/1% BME/0.5% sarkosyl) for 20 minutes. Arrays were then incubated in a hybridization buffer (2M NaCl/8% v/v PEG8000) for 40 minutes, and then the beads were removed from the arrays and collected in 1.5 mL tubes in wash buffer (2M NaCl/3 mM MgCl2/20 mM Tris-HCl/8% v/v PEG8000). Beads were resuspended in a reverse transcription master mix, and reverse transcription, exonuclease digestion, second strand synthesis, and whole transcriptome amplification were carried out as previously described. Libraries were generated using Illumina Nextera XT Library Prep Kits and sequenced on NextSeq 500/550 High Output v2.5 kits to an average depth of 110 million reads per array: read 1: 21 (cell barcode, UMI), read 2: 50 (digital gene expression), index 1: 8 (N700 barcode).

### Single-Cell Computational Pipelines, Analysis & Annotation

#### Single-Cell RNA-seq Computational Pipelines, Processing, and Analysis (Columbia)

We pseudo-aligned the raw reads for each sample to a merged human/SARS-CoV-2 transcriptome using kallisto v0.46.2 in “BUS” mode to facilitate demultiplexing (Bray et al. 2016; *Melsted et al. 2019*). We then corrected for index swapping across all samples that were sequenced in a given lane at the level of equivalence classes using the algorithm of Griffiths et al. (*Griffiths et al. 2018*). Finally, we generated a raw count matrix based on the index swap-corrected BUS file for each sample using bustools v0.40.0 (*Melstad et al. 2019*). We filtered the raw count matrix for each sample using the EmptyDrops algorithm from Lun et al (*Lun et al. 2019*) and also removed all cells with mitochondrial pseudo-alignment rates >20% or with counts per gene greater than two standard deviations above the mean for a given sample.

For preliminary cell type annotation, we first analyzed each sample individually. We identified likely markers of cell types and subpopulations based on the drop-out curve for each sample as described in Levitin et al. (*Levitin et al., 2019*) We then merged all of the count matrices for airway and, separately, all of the count matrices for blood for cell type annotation. We performed unsupervised clustering on the merged airway and blood matrices by taking the union of putative marker genes identified from each sample, creating a submatrix for putative marker genes, and transforming this into a matrix of Spearman correlation distances. We then used this distance matrix as input for unsupervised clustering with Louvain community detection as implemented in Phenograph with k=20 for k-nearest neighbor graph generation (*Levine et al. 2015*). We identified specific marker genes for each cluster using the binomial test as described in Shekhar et al.(*Shekhar et al. 2016*) The Python code used for this analysis can be found at https://github.com/simslab/cluster_diffex2018.

### Single-Cell RNA-seq Computational Pipelines and Analysis (Sanger & IJC)

The single cell transcriptome and CITEseq quantification was performed using cellranger 3.1, using GRCh38 + SARSCoV2 reference and a dictionary of tagged antibodies tags respectively. Several i7 sample tags were used for pairs of donors for the PBMCs samples, so the provenance of such cells was resolved using Souporcell (*Du et al. 2020*). This tool produced a genotype variant for every donor that could be matched to genotyping obtained from the Infinium Global Screening Array-24 v3.0 BeadChip, and a list of cell doublet was resolved from observing features that cannot be explained by a single genotype. In addition, Scrublet (*Wolock et al. 2019*) was used to detect other doublets by detecting cell as outliers for their computed doublet score, by using a t-test with associate pvalue less than 0.01 after Bonferroni correction within fine-grained sub-clustering of each cluster produced by the Leiden algorithm from ScanPy (*Wolf et al. 2018)*, which was performed separately for PMBCs and nasal swabs. Cells detected as doublet by either method and other cells that have a high mitochondrial content (>15%) were excluded from downstream analysis.

The cell type identities were resolved using scVI (*Lopez et al. 2018*) which only used transcriptome information. Sample specific batch effects were corrected by a pair of generative models. For the PBMCs samples, the correction was performed using both cell-wise deconvolved donor identities and sequencing lane, which resulted in one batch for every 13 PBMcs samples and 13 additional batches for their respective CD3N sorted counterparts. The second generative model would be trained on the 4 nasal swab samples, which was processed separately as 4 batches. Genes associated with G1 and S cell stage, which are provided in the Seurat package (*Stuart et al. 2019*), were excluded and the remaining genes were downsampled to 5000 genes using scvi native method. The number of latent variables needed to infer had been set of 64, and the associated generative models were for PMBCs and nasal swabs trained separately using 500 iterations.

Scanpy (*Wolf et al. 2018*) was then used on the inferred latent variables to identify cell-type clusters and render umap coordinates that would be available to browser using cellxgene. Sub-clustering of the Bcell population and Tcell populations were performed for the PBMCs sample in order to resolve expected cell-types that were not directly discriminated by the unsupervised analysis. For instance, the naive Tcell populations could be resolved from high level of CCR7, and also from having levels of antibody_CD45RA as opposed to antibody_CD45RO, which is high in other CD4+ Tcells. Similarly, memory Bcells were identified from having higher levels for CD27.

#### Data Preprocessing and Quality Control

Pooled libraries were demultiplexed using bcl2fastq (v2.17.1.14) with default settings (mask_short_adapter_reads 10, minimum_trimmed_read_length 10). Libraries were aligned using STAR within the Drop-Seq Computational Protocol (https://github.com/broadinstitute/Drop-seq) and implemented on Cumulus (https://cumulus.readthedocs.io/en/latest/drop_seq.html). Reads were aligned to a custom combined genome of human GRCh38 (Ensembl 93) and SARS-CoV-2 RNA genome. The SARS-CoV-2 viral sequence and gtf are as described in Kim et al. 2020 (https://github.com/hyeshik/sars-cov-2-transcriptome, BetaCov/South Korea/KCDC03/2020 based on NC_045512.2). The GTF was edited to include all CDS regions (as of this annotation of the transcriptome, the CDS regions completely cover the RNA genome without overlapping segments), and regions were added to describe the 5’ UTR (“SARSCoV2_5prime”), the 3’ UTR (“SARSCoV2_3prime”), and reads aligning to anywhere within the Negative Strand (“SARSCoV2_NegStrand”). Additionally, trailing A’s at the 3’ end of the virus were excluded from the SARS-CoV-2 fasta, as these were found to drive spurious viral alignment in pre-COVID19 samples. Alignment references were tested against a diverse set of pre-COVID-19 samples and *in vitro* SARS-CoV-2 infected human bronchial epithelial cultures (Ravindra et al.) to confirm specificity of viral aligning reads (data not shown). Aligned cell-by-gene matrices were merged across all study participants, and cells were filtered to eliminate barcodes with fewer than 200 UMI, 150 unique genes, and greater than 50% mitochondrial reads (cutoffs determined by distributions of reads across cells). A preliminary clustering analysis as described below was carried out, which identified a subset of cells defined solely by high mitochondrial alignment, and these cells were removed. As of 07/29/2020, this resulted in an interim dataset of 32,389 genes and 12,427 cells across 21 study participants (15 COVID-19 individuals, 6 control individuals) with a mean +/- SEM recovery per swab of 565 +/- 123 cells. 1 additional nasal swab was processed and sequenced, but excluded from analysis as no high-quality cell barcodes were recovered after sequencing (NB: this sample contained < 5,000 viable cells prior to loading).

#### Cell Clustering and Annotation

Dimensionality reduction, cell clustering and differential gene analysis were all achieved using the Seurat (v3.1.5) package in R programming language (v3.0.2). Dimensionality reduction was carried out by running principal components analysis over the 2,291 most variable genes with dispersion > 0.9 (tested over a range of dispersion > 0.7 to dispersion > 1.2; dispersion > 0.9 was determined as optimal based on number of variable genes, and general stability of clustering results across these cutoffs was confirmed). Only variable genes from human transcripts were considered for dimensionality reduction and clustering. Using the Jackstraw function within Seurat, we selected a subset of 24 principal components that described the majority of variance within the dataset, and used these for defining a nearest neighbor graph and Uniform Manifold Approximation and Projection (UMAP) plot. Cells were clustered using Louvain clustering, and the resolution parameter was chosen by maximizing the average silhouette score across all clusters. Differentially expressed genes between each cluster and all other cells were calculated using the FindAllMarkers function, test.use set to “bimod”. After initial clustering, we pooled all clusters putatively determined to be of epithelial origin and reanalyzed this subset to find subclusters, using the methods for dimensionality reduction and clustering as above (dispersion cutoff > 0.9, 25 principal components). Within the epithelial cell subcluster, we then further subclustered cells belonging to the ciliated cell, squamous cell, and secretory cell Louvain clusters separately, resolving specific subtypes and substates among each cell type. Similar to epithelial cells, we combined all Louvain clusters containing myeloid cell types and reanalyzed as above (dispersion > 1, mean expression > 0.3, 6 principal components). After complete subclustering, all cell type identities were combined together to generate an interim annotated dataset.

### Single-Cell RNA-seq Computational Pipelines, Processing, and Analysis (VIB/University Hospital Ghent)

The raw reads were demultiplexed and mapped to a merged human/SARS-CoV-2 genome using Cell Ranger v4.0. Empty droplets and outlier cells were identified and removed based on the gene expression profile. Cells with counts in less than 200 genes and genes expressed in less than 3 cells were removed from the count matrix. Cells that were more than 5 mean absolute deviations from the median library size or median number of expressed genes were also removed, as well as cells where the % of mitochondrial reads exceeded the median by 5 mean absolute deviations. The ensuing count matrix was further processed using Seurat v3.1.5. The gene expression counts were divided by the library size and after applying a scaling factor log-transformed to normalize between cells. A centered log-ratio transform was used to normalize the antibody derived counts. Cells were clustered using the Louvain algorithm on the 50 first principal components of a subset of high variable genes and visualized on a Uniform Manifold Approximation and Projection of a batch-effect corrected embedding using Harmony (*Korsunsky et al., 2019*). The computational resources (Stevin Supercomputer Infrastructure) and services were provided by the VSC (Flemish Supercomputer Center), funded by Ghent University, FWO and the Flemish Government – department EWI.

### Single-Cell RNA-seq Computational Pipelines, Processing, and Analysis (Sanger/UCL)

The single cell data was mapped to a GRCh38 ENSEMBL 93 derived reference, with an additional 21 viral genomes (featuring SARS-CoV2) included as additional FASTA sequences and corresponding GTF entries. A complete list of the included viruses, along with their respective NCBI IDs and source links, can be found in Supplementary Table X. The alignment, quantification and preliminary cell calling were carried out via the STARsolo functionality of STAR 2.7.3a, with the cell calling subsequently refined with Cell Ranger 3.0.2’s version of EmptyDrops (Lun et al., 2019). This algorithm has been made available as emptydrops on PyPi. Initial doublets were called on a per-sample basis by computing Scrublet (*Wolock et al., 2019*) scores for each cell, propagating them through an overclustered manifold by replacing individual scores with per-cluster medians, and identifying statistically significant values from the resulting distribution, replicating the approach of *Pijuan-Sala et al. 2019* and *Popescu et al. 2019*. The clustering was performed with the Leiden (*Traag et al. 2019*) algorithm on a KNN graph of a PCA space derived from a log(CPM/100 + 1) representation of highly variable genes, following SCANPY protocol (*Wolf et al. 2018*), and overclustering was achieved by performing an additional clustering of each resulting cluster. The primary clustering also served as input for ambient RNA removal via SoupX (*Young et al. 2020*).

### Single-Cell RNA-seq Computational Pipelines, Processing, and Analysis (MGH & Broad; Acute Blood Cohort PBMC Analysis)

We ran Cell Ranger 3.1.0 (10x Genomics) on Terra (Broad Institute) using the cellranger_workflow WDL file included in Cumulus (*Li et al. 2020*). This pipeline runs the “cellranger mkfastq’’ command to demultiplex raw sequencing reads, and the “cellranger count” command to align the sequencing reads and generate a counts matrix. We aligned to a custom-built Human GRCh38 (Ensembl 93) and SARS-COV-2 RNA reference. The SARS-CoV-2 viral sequence and GTF are as described in *Kim et al. 2020* (https://github.com/hyeshik/sars-cov-2-transcriptome, BetaCov/South Korea/KCDC03/2020 based on NC_045512.2). The GTF was edited to include all CDS regions (as of this annotation of the transcriptome, the CDS regions completely cover the RNA genome without overlapping segments), and regions were added to describe the 5’ UTR (“SARSCoV2_5prime”), the 3’ UTR (“SARSCoV2_3prime”), and reads aligning to anywhere within the Negative Strand (“SARSCoV2_NegStrand”). Additionally, trailing A’s at the 3’ end of the virus were excluded from the SARS-CoV-2 fasta, as these were found to drive spurious viral alignment in pre-COVID19 samples. PBMC samples were pooled by multiplexing 8 samples. Donor identity was deconvolved using demuxEM on the hashtagged samples (*Gaublomme et al. 2019*).

#### Performing quality control, data integration of individual samples and clustering analysis

Single-cell RNA-seq data from individual samples were combined into a single large gene expression matrix and analyzed using R (https://www.R-project.org). Cells were removed in each sample if they met the following criteria:

- <100 UMIs / cell
- <500 genes detected / cell
- >20% mitochondrial UMIs / cell
- <0.01% mitochondrial UMIs / cell

We selected genes with high variation by fitting a Loess regression (log(sd) ∼ log(mean)) to the 13,357 genes in the count data with expression in at least 300 cells. We took the 2599 genes with greatest residual variance from the Loess fit, and ran principal component analysis (PCA) on these scaled and centered genes to compute the top 30 principal components. To adjust each of the PCs, we ran Harmony (*Korsunsky et al. 2019*) for 25 iterations on the 30 PCs, where each multiplexed sample was considered its own batch. Next, we used the 30 adjusted PCs to compute the 50 nearest neighbors (Euclidean distance) for each cell with the HNSW algorithm (*Malkov and Yashunin 2016*) as implemented in the BiocNeighbors R package. The network of nearest neighbors was used as the input for unsupervised clustering with the Leiden community detection algorithm (*Traag et al. 2018*) as implemented in the leidenalg Python package (https://github.com/vtraag/leidenalg) with resolution set to 0.8 for 20 iterations. To embed the cells in a two-dimensional map, we ran UMAP on the nearest neighbor network as implemented in the uwot R package (https://github.com/jlmelville/uwot) with spread set to 1 and min_dist set to 0.25.

For preliminary cell type annotation, we identified specific marker genes for each cluster using the area under the receiver operator curve (AUROC) computed by the Presto R package (https://github.com/immunogenomics/presto). We also used the p-value from the linear model fit (gene ∼ in_cluster) on pseudobulk expression (*Lun et al. 2016*) using the Limma R package (*Ritchie et al. 2005*).

### Ethics Declarations

- Columbia
  ○ Full name of Ethics Committee / Institutional Review Board (IRB) for the cohort: Columbia University Irving Medical Center
  ○ Decision made by ethical oversight body: Approved IRB protocol AAAS9659
- UMMC/BCH/Broad
  ○ Full name of Ethics Committee / Institutional Review Board (IRB) for the cohort: University of Mississippi Medical Center
  ○ Decision made by ethical oversight body: Approved UMMC IRB#2020-0065
- MGH/Broad
  ○ Full name of Ethics Committee / Institutional Review Board (IRB) for the cohort: Partners Human Research Committee (now renamed Mass General Brigham IRB)
  ○ Decision made by ethical oversight body: The addition of the COVID cohort to 2107P001681 was approved via amendment with a waiver of informed consent for this cohort.
- Wellcome Sanger Institute/Josep Carreras Research Institute (IJC)
  ○ Full name of Ethics Committee / Institutional Review Board (IRB) for the cohort: Germans Trias i Pujol Hospital
  ○ Decision made by ethical oversight body: Approved IRB protocol PI-20-129
  ○ Full name of Ethics Committee / Institutional Review Board (IRB) for the cohort: Bellvitge Hospital Universitari
  ○ Decision made by ethical oversight body: Approved IRB protocol AC010/20
  ○ Full name of Ethics Committee / Institutional Review Board (IRB) for the cohort: Hospital Universitari Vall d’Hebron
  ○ Decision made by ethical oversight body: Approved IRB protocol PR(AG)282/2020
  ○ Full name of Ethics Committee / Institutional Review Board (IRB) for the cohort: Hospital Universitario de la Princesa
  ○ Decision made by ethical oversight body: Approved IRB protocol 4070
  ○ Full name of Ethics Committee / Institutional Review Board (IRB) for the cohort: Universitätsklinikun Freiburg
  ○ Decision made by ethical oversight body: Approved IRB protocol 507/16; 282/11
- VIB/Ghent University Hospital
  ○ Full name of Ethics Committee / Institutional Review Board (IRB) for the cohort: Ethics Committee of Ghent University Hospital (Belgium), AZ Jan Palfijn (Belgium) and AZ Maria Middelares (Belgium)
  ○ Decision made by ethical oversight body: approved reference G0G4520N.
- Wellcome Sanger Institute/University College London
  ○ Full name of Ethics Committee / Institutional Review Board (IRB) for the cohort: Living Airway Biobank, administered through UCL Great Ormond Street Institute of Child Health
  ○ Decision made by ethical oversight body: Approved REC reference: 19/NW/0171, IRAS project ID 261511

## Data Availability

Processed data is openly and freely available to explore via the covid19cellatlas.org website. The underlying processed data is also openly and freely available for download and reuse with requested attention to the community standards that the authors outline. Data will be updated and shared going forward to encourage prepublication reuse.

## Full Author Table

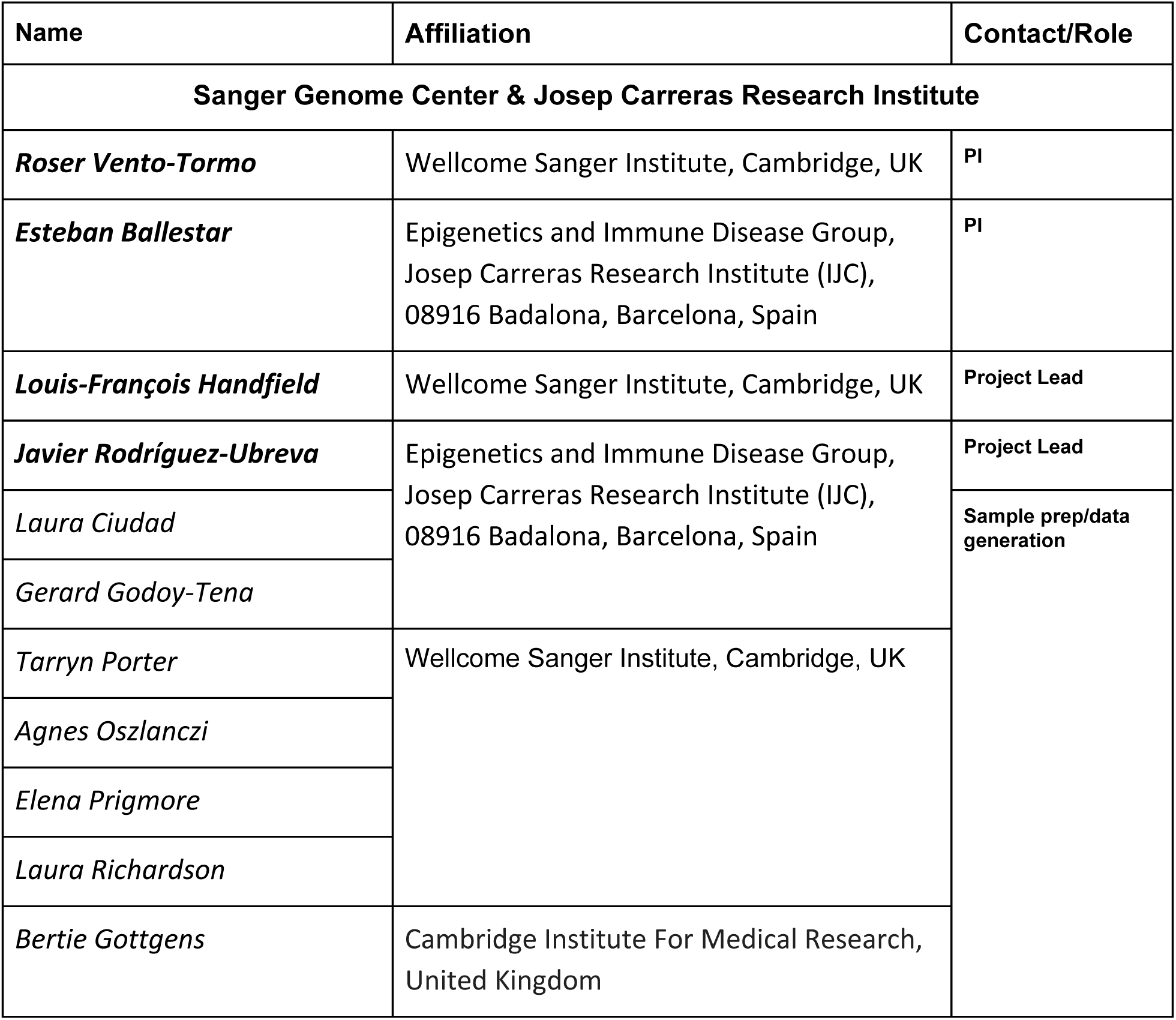
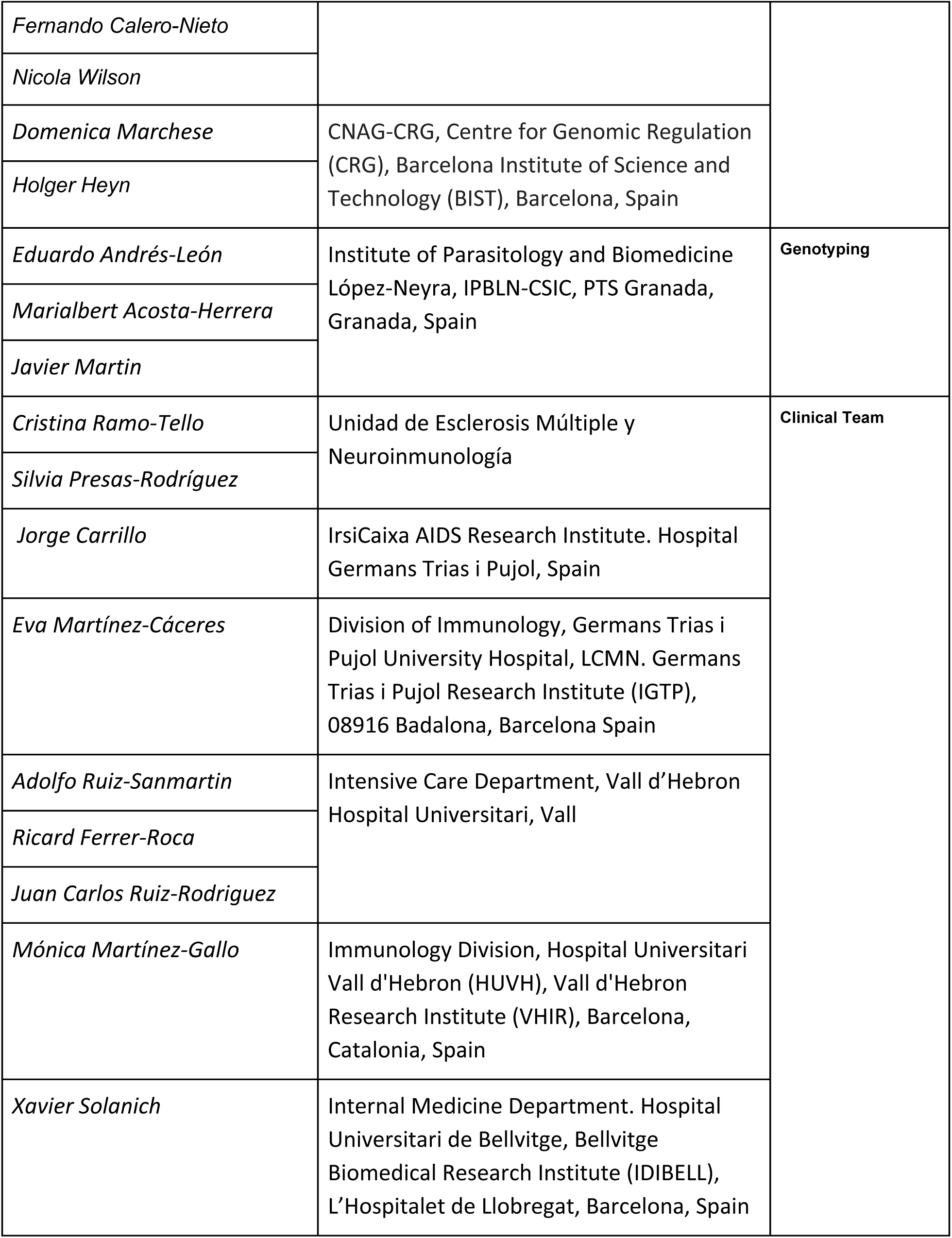
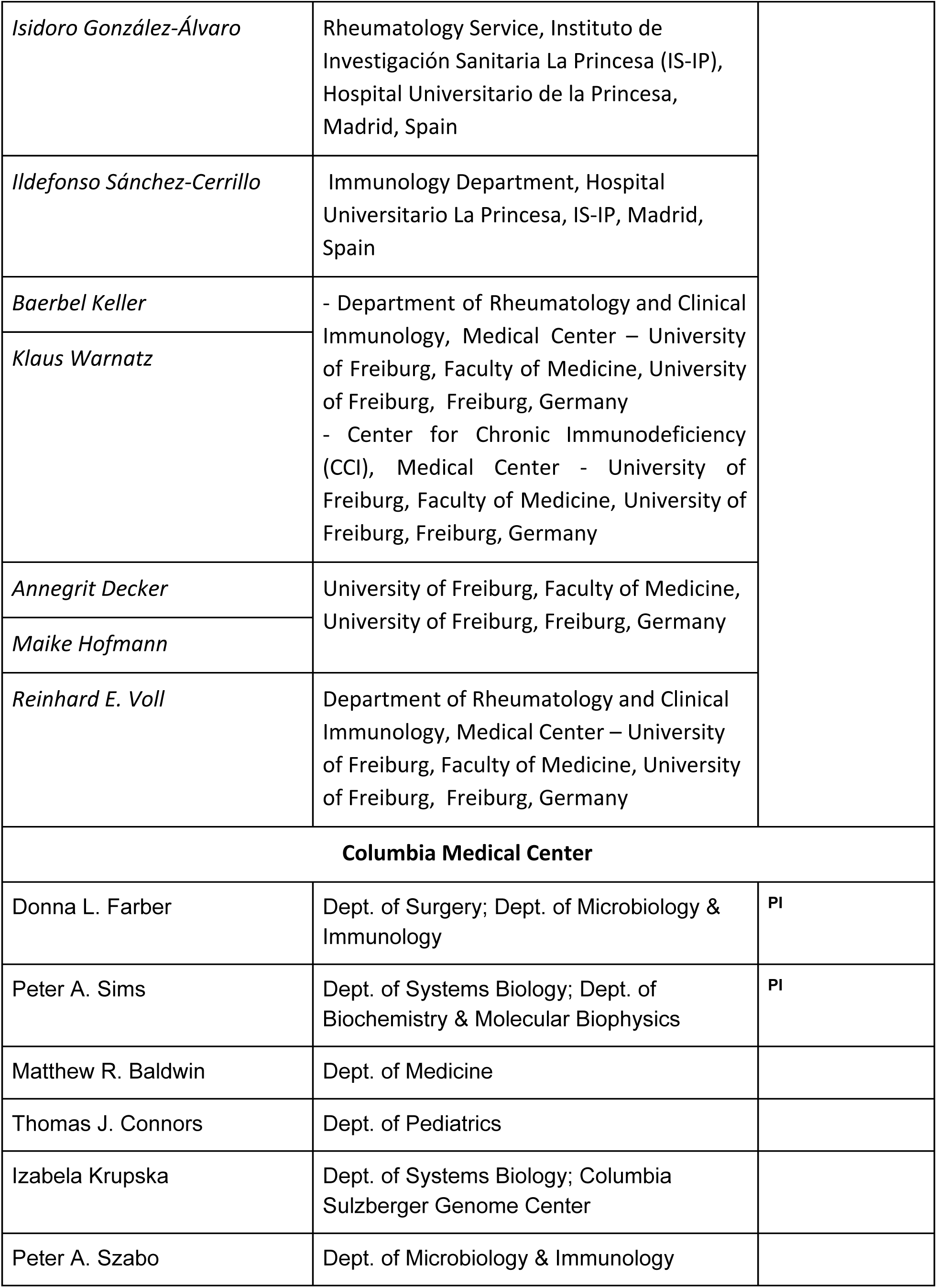
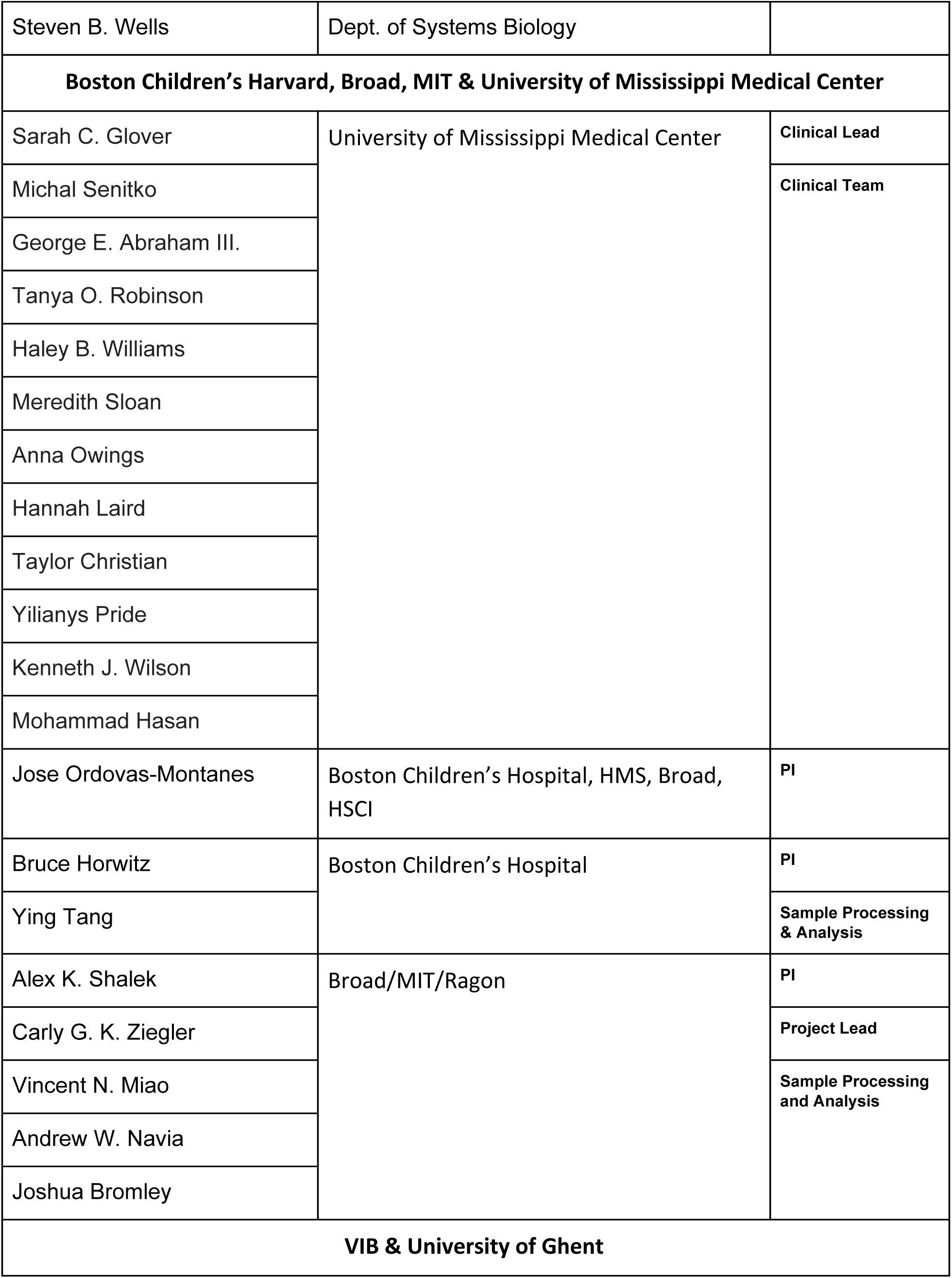
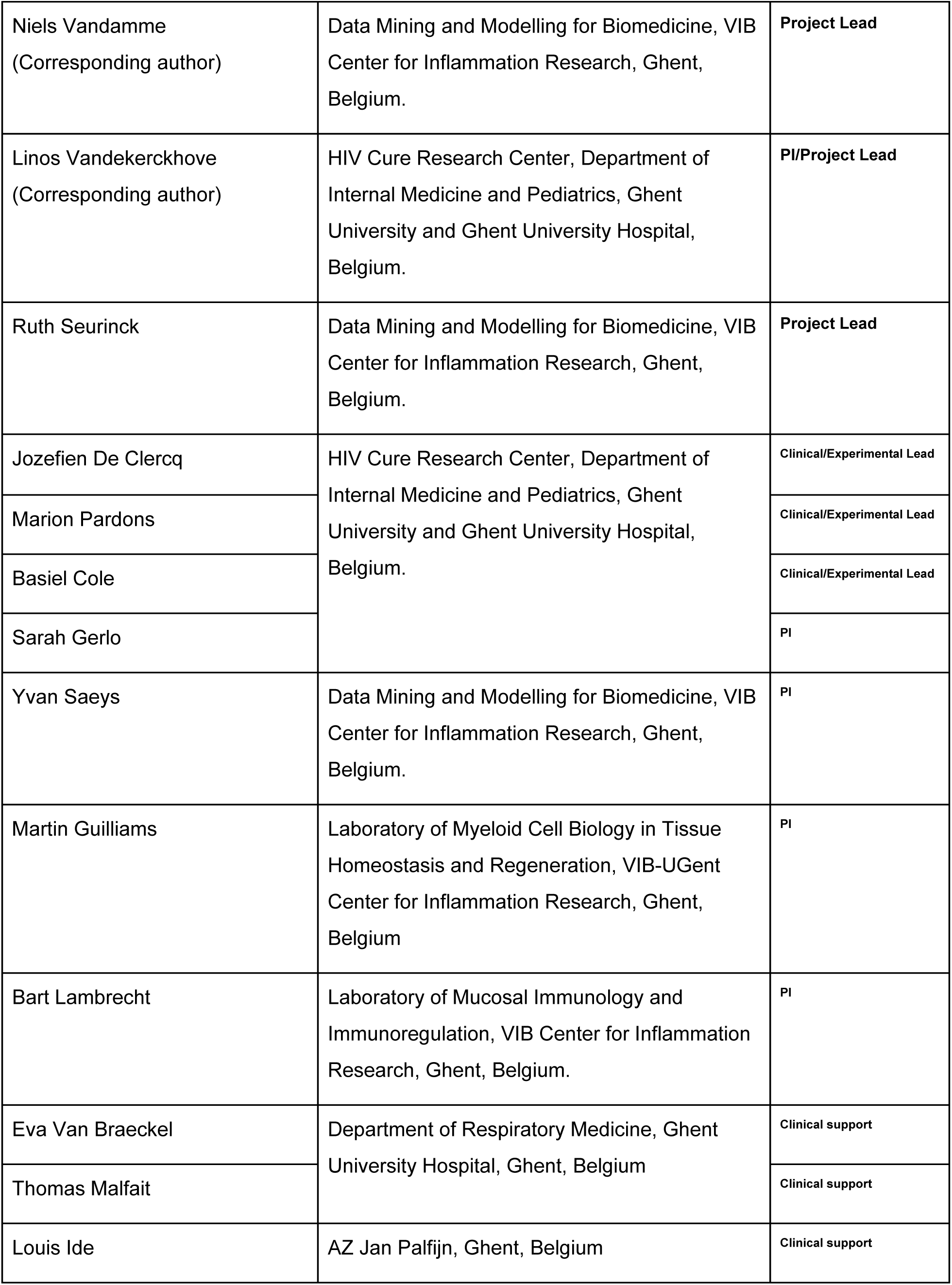
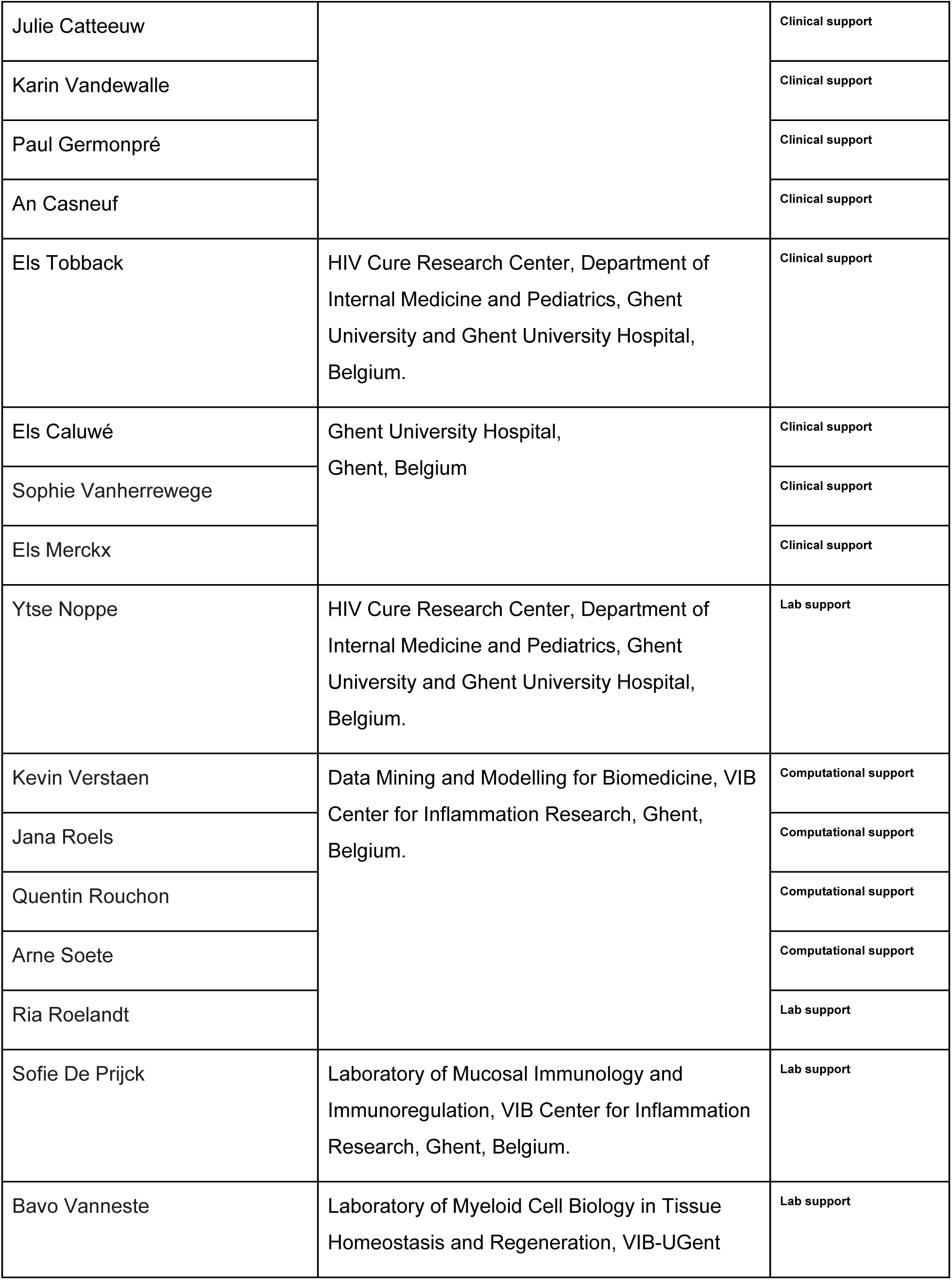
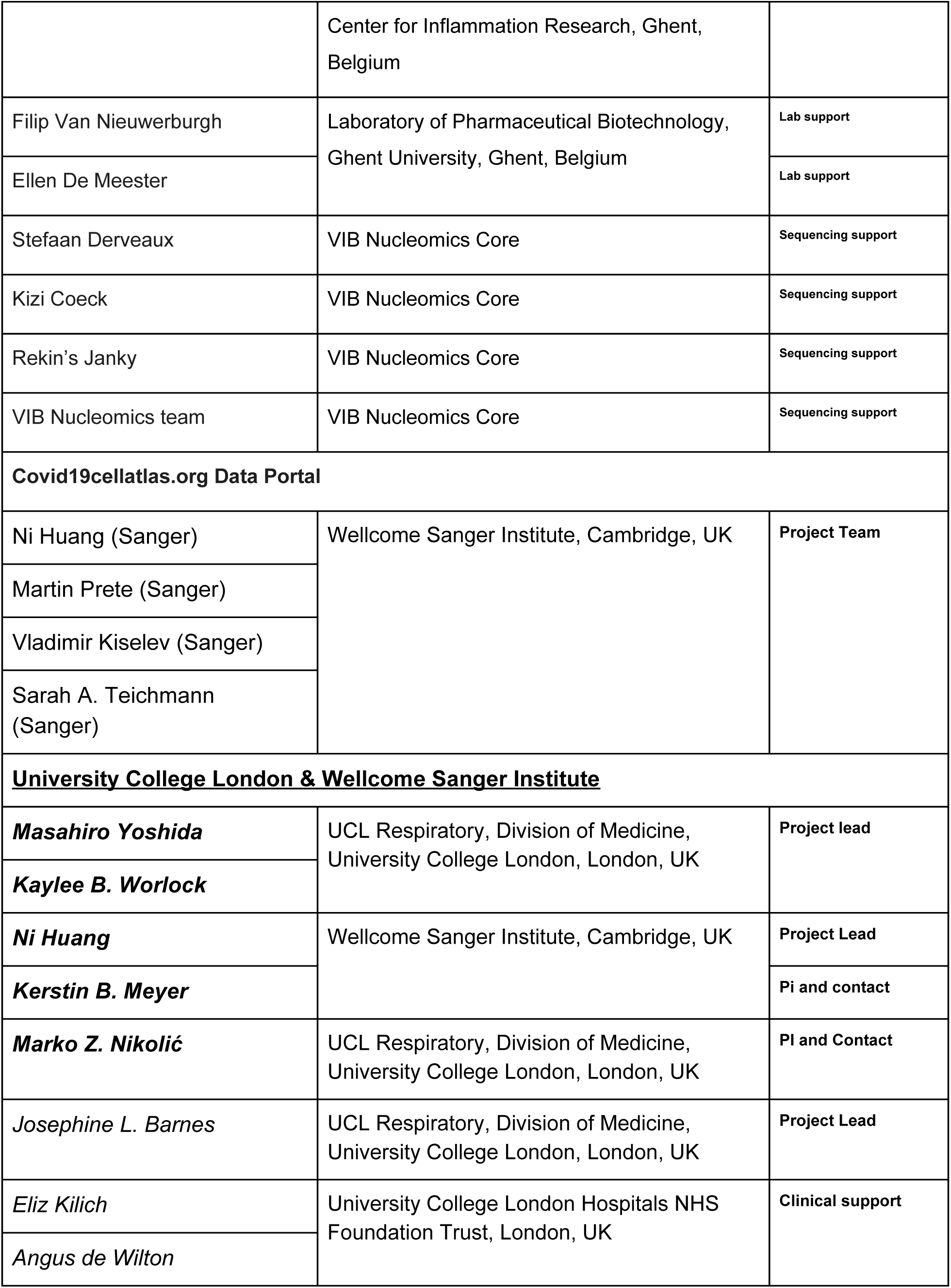
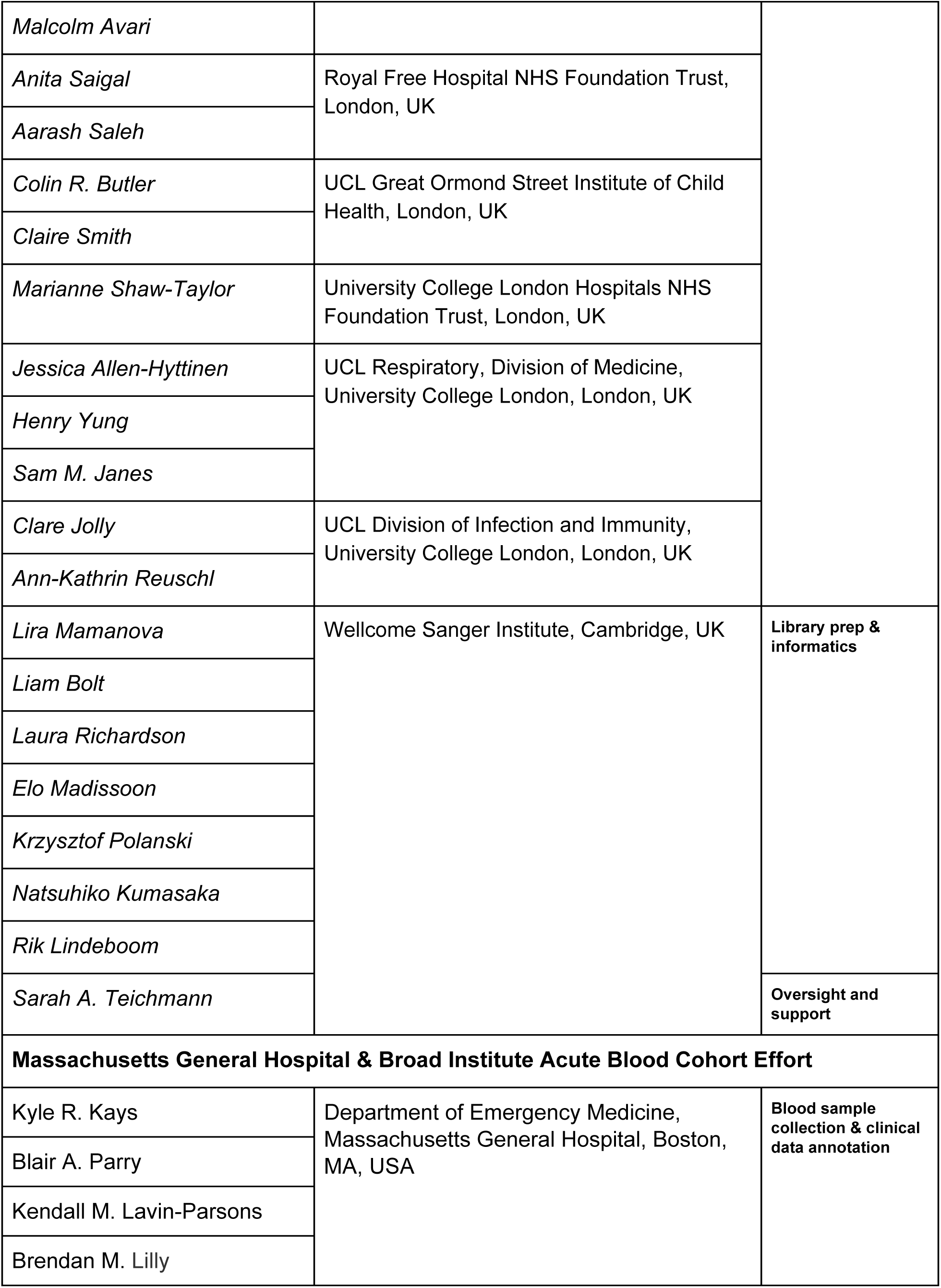
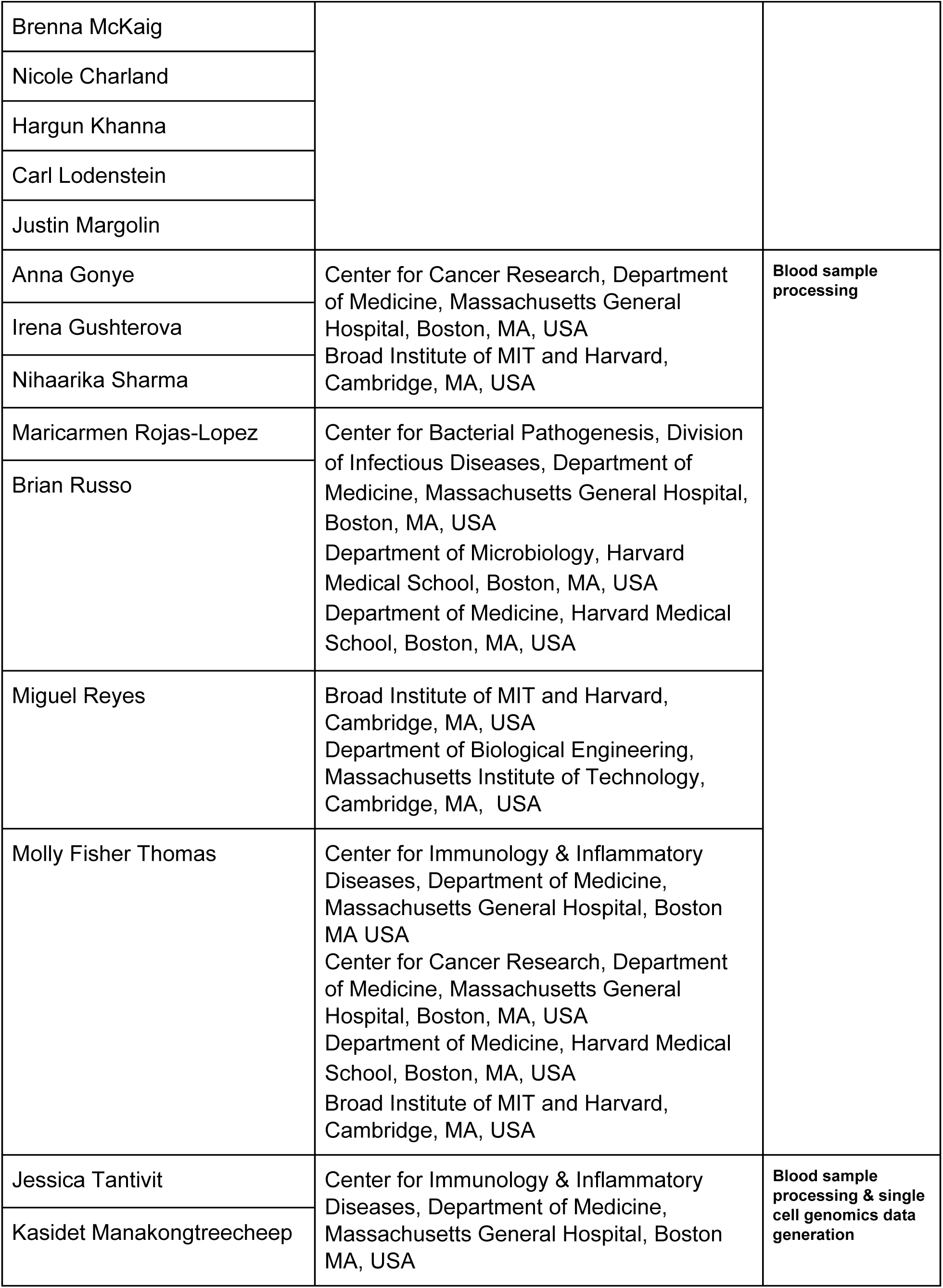
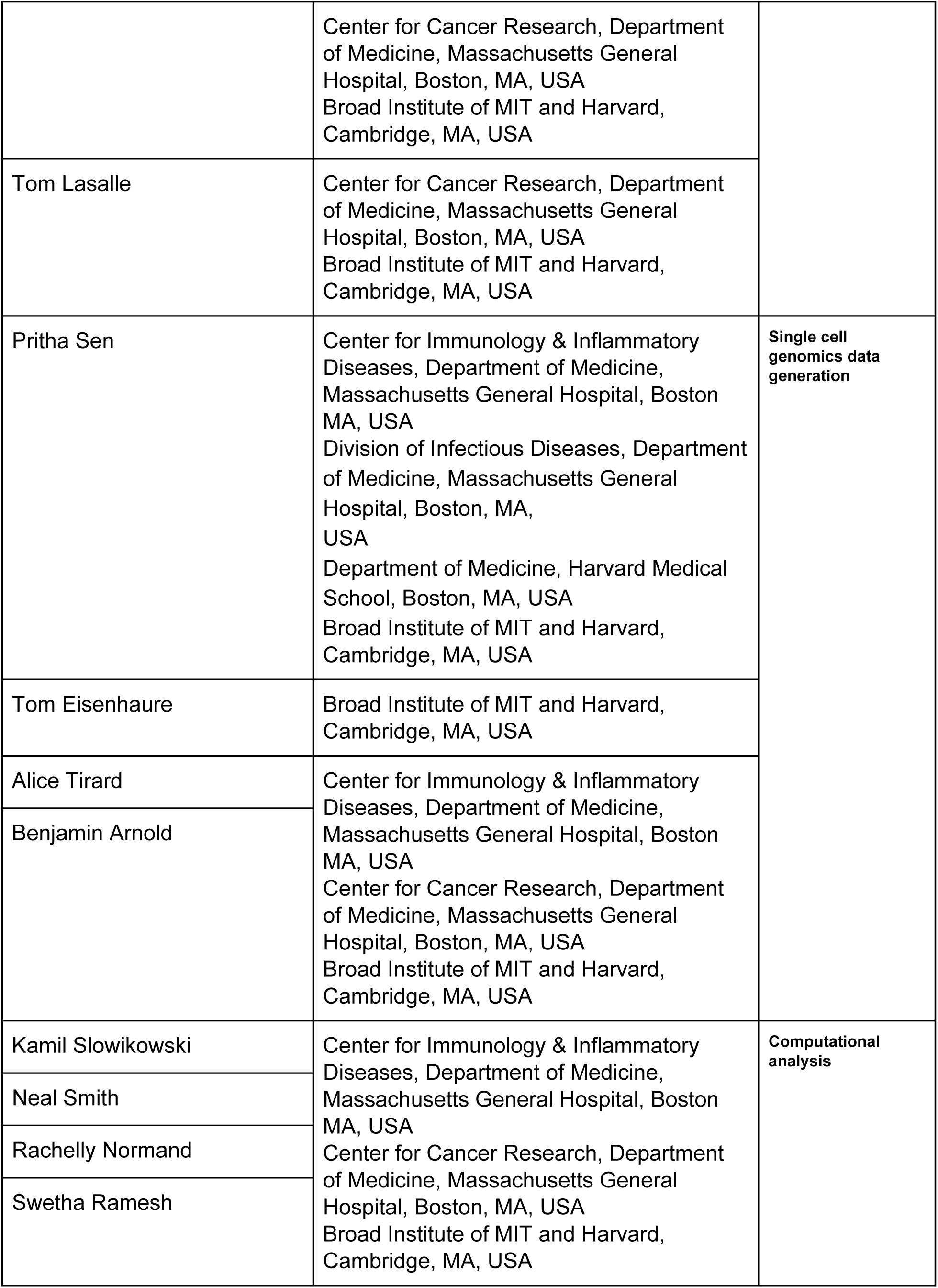
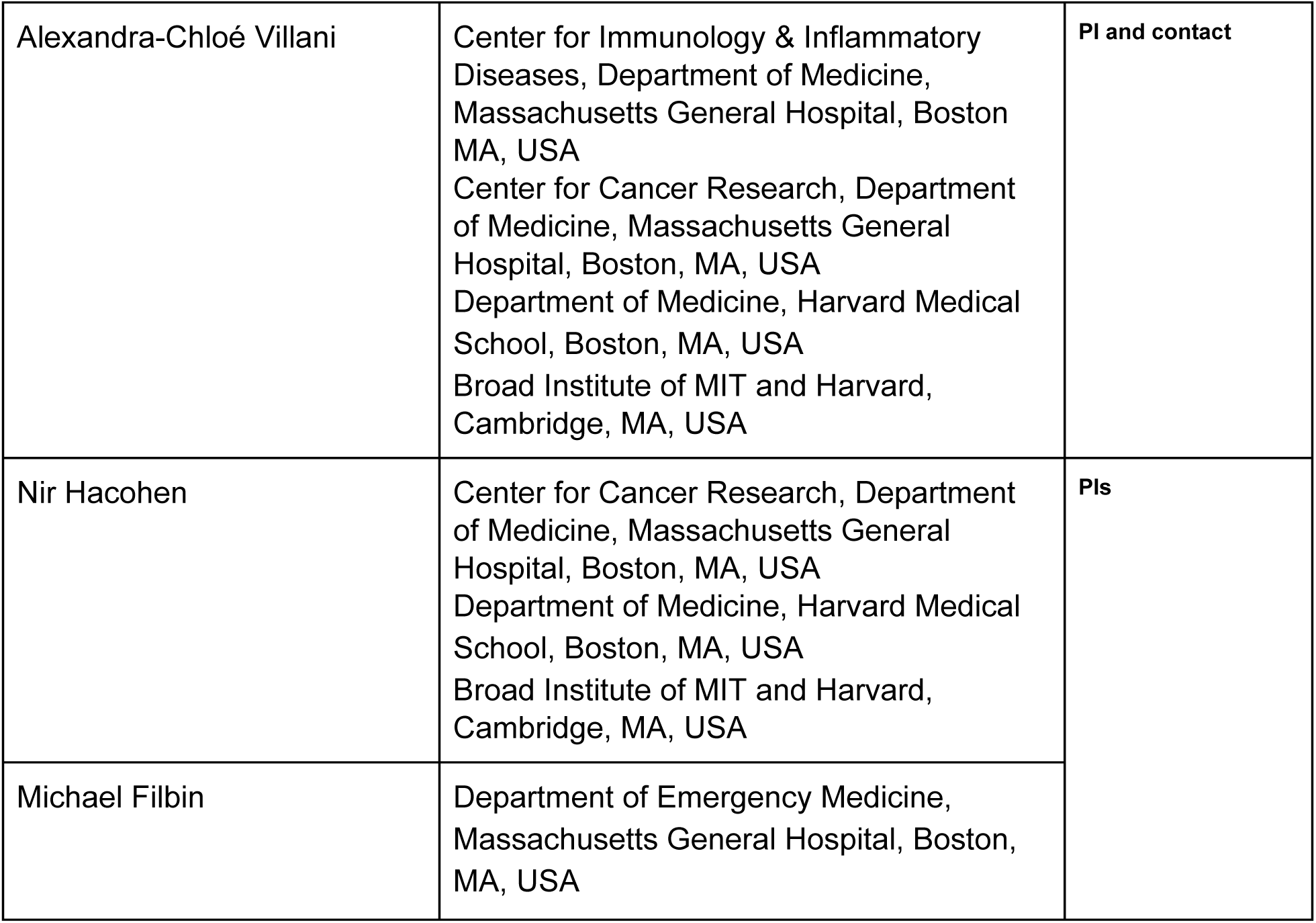
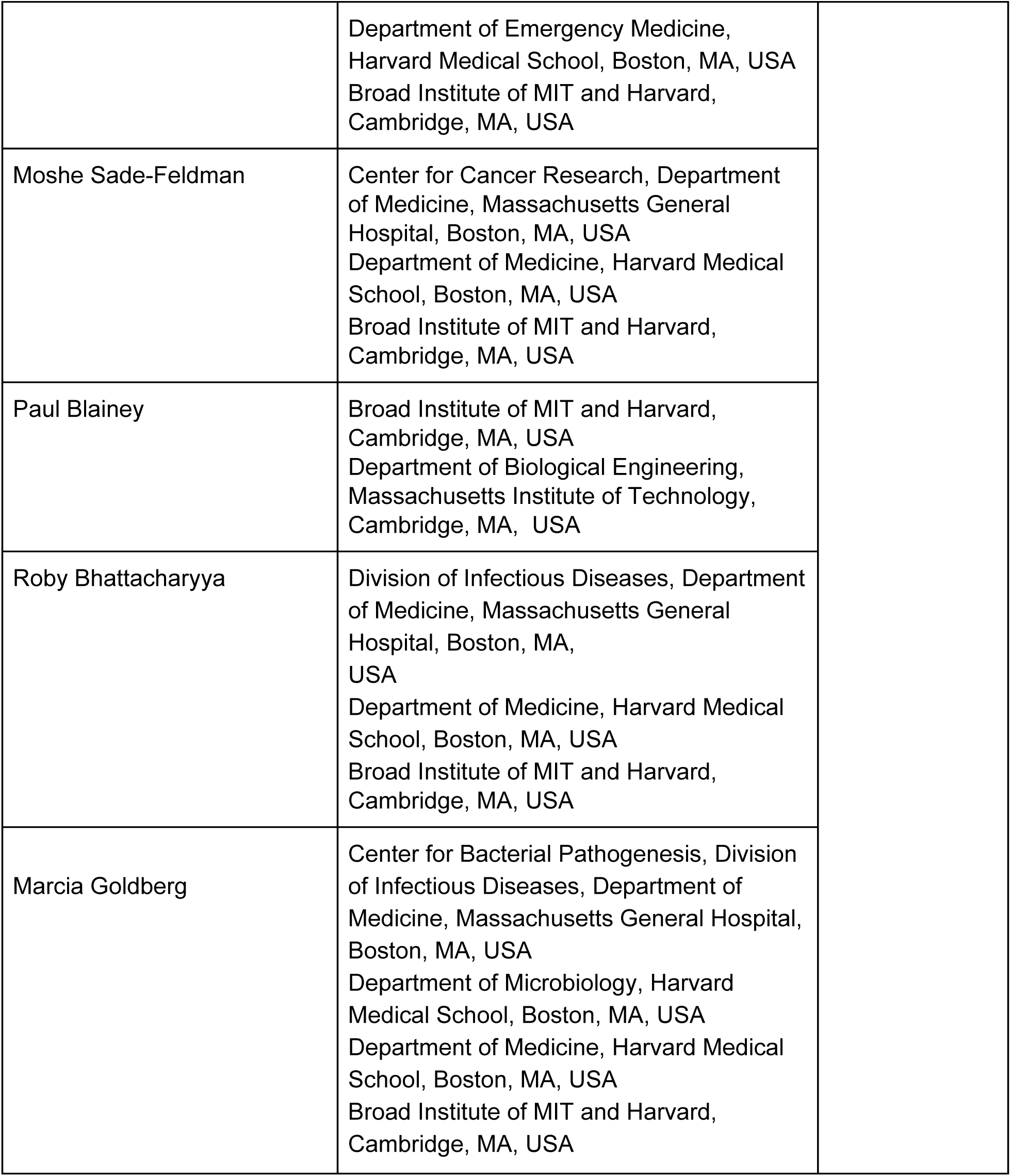

